# An immune dysfunction score for stratification of patients with acute infection based on whole blood gene expression

**DOI:** 10.1101/2022.03.17.22272427

**Authors:** Eddie Cano-Gamez, Katie L Burnham, Cyndi Goh, Zunaira H Malick, Andrew Kwok, David A Smith, Hessel Peters-Sengers, David Antcliffe, GAinS Investigators, Stuart McKechnie, Brendon P Scicluna, Tom van der Poll, Anthony C Gordon, Charles J Hinds, Emma E Davenport, Julian C Knight

## Abstract

Dysregulated host responses to infection can lead to organ dysfunction and sepsis, causing millions of deaths globally each year. To alleviate this burden, improved prognostication and biomarkers of response are urgently needed. We investigated the use of whole blood transcriptomics for stratification of patients with severe infection by integrating data from 3,149 samples of sepsis patients and healthy individuals into a gene expression reference map. We used this map to derive a quantitative sepsis response signature (SRSq) score reflective of immune dysfunction and predictive of clinical outcomes, which can be estimated using a 19-gene signature. Finally, we built a machine learning framework, SepstratifieR, to deploy SRSq in sepsis, H1N1 influenza, and COVID-19, demonstrating clinically relevant stratification across diseases and revealing the physiological alterations linking immune dysregulation to mortality. Our method enables early identification of individuals with dysfunctional immune profiles, thus bringing us closer to precision medicine in infection.

## Introduction

Infectious diseases result in considerable global morbidity and mortality.^1^ Notably, the recent H1N1 influenza^2^ and SARS-CoV-2^3^ pandemics illustrate how individuals without significant risk factors can still develop critical illness following infection. In extreme cases, this can lead to sepsis, a dysregulated host response accompanied by major organ dysfunction^4^ which accounted for 11 million deaths in 2017 alone.^5^ This highlights the importance of better understanding maladaptive host immune responses. In particular, it is fundamental to recognise which patient immune profiles are dysfunctional and amenable to interventions like immunomodulatory therapy or early organ support.

High-throughput technologies can group individuals by molecular profile, thus enabling patient stratification.^6^ In sepsis, a number of patient subgroups (i.e. endotypes) haven been proposed based on whole blood leukocyte gene expression.^7–11^ We previously described two sepsis response signature (SRS) groups: SRS1, an immunocompromised profile showing increased risk of death, and SRS2, characterised by immunocompetency and reduced mortality.^7^ These also showed differential responses to corticosteroid treatment.^12^ Nevertheless, similar developments are lacking for the wider population of patients with infection, who do not always fulfil conventional sepsis criteria. Stratification methods applicable across multiple infecting pathogens, independently of severity or technological differences, are vital. Ideally, these should be used in conjunction with technologies with rapid turn-around suitable for point-of-care testing.^13,14^

We developed SepstratifieR, a machine learning framework which addresses these limitations. SepstratifieR was trained on data from 3,149 samples of sepsis patients and healthy volunteers encompassing three technological platforms. This makes it a highly flexible framework which is largely independent of technological differences, amenable to point-of-care testing, and applicable to a broad range of infections. SepstratifieR achieves personalised risk prediction by deriving a score reflective of each patient’s level of immune dysfunction. We show that this score can model disease heterogeneity more accurately and advance outcome prediction. Finally, we demonstrate that SepstartifieR achieves clinically meaningful stratification in bacterial/viral sepsis, influenza, and COVID-19.

## Results

### A cross-platform transcriptional map of the host response in sepsis

Sepsis endotypes have been extensively described using microarrays.^7–10^ However, it is unclear if they generalise to sequencing-based assays. We assessed whether our previously proposed SRS endotypes^7^ were detectable using RNA-seq by leveraging data from 134 patients from the UK Genomic Advances in Sepsis (GAinS) study with both microarray and RNA-seq measurements available. We used canonical correlation analysis (CCA) to create a joint representation of both assays (**Methods**).^15^ In brief, CCA identifies linear combinations of variables (canonical dimensions) which maximise the correlation between two data sets, representing shared axes of variation. Based on labels known from previous studies^7,8^, we demonstrated that the first canonical dimension (CC1) separated SRS1 from SRS2 patients (**Figure 1A**), suggesting that SRS endotypes are identifiable using RNA-seq.

**Figure 1.**
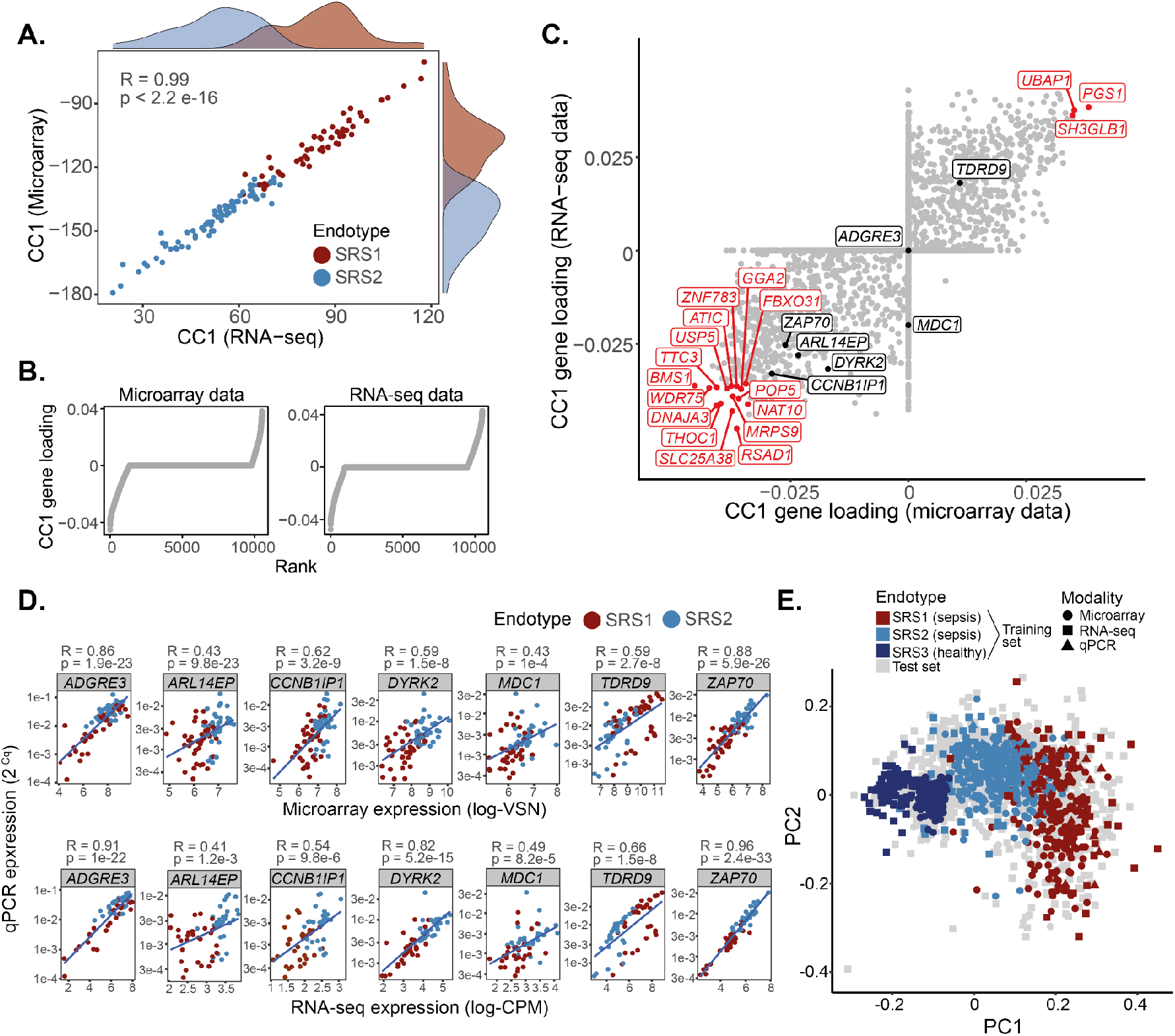
Construction of a reference map of gene expression in sepsis using data from three different platforms. **A)** Sparse canonical correlation analysis of samples with RNA-seq and microarray measurements in the GAinS study. Each dot represents a sample, with X and Y coordinates corresponding to the first canonical component as estimated from RNA-seq and microarray data, respectively. Marginal distributions are provided, with each histogram representing the distribution of either SRS1 (red) or SRS2 (blue) samples along the two canonical components. R = Pearson correlation coefficient; p = correlation p value estimated using a two-tailed T test. **B)** Contribution of each gene to the first canonical component (Y axis; i.e. gene loading) as estimated from microarray and RNA-seq data, respectively. Dots represent genes, ranked by increasing CC1 loading (X axis). **C)** Scatter plot of CC1 gene loadings for each gene as calculated from microarray (X axis) and RNA-seq (Y axis) data. Each dot represents a gene. Black dots indicate genes in the Davenport signature. Red genes represent the top 1% genes with highest CC1 contribution in both assays. **D)** Correlation of gene expression measurements between microarray or RNA-seq (X axis) and qRT-PCR (Y axis) for all genes in the Davenport signature. Each dot represents a sample, with colours indicating its respective sepsis endotype. Lines represent the best linear fit. R = Pearson correlation coefficient; p = correlation p value estimated using a two-tailed T test. **E)** Reference map of gene expression in sepsis constructed based on the Davenport signature. Data from four different cohorts were integrated using mNN batch correction, followed by visualisation using PCA. Each dot represents a sample, with colours indicating its corresponding sepsis endotype. Grey dots represent samples in the test set, for which endotype is not known. Different shapes indicate the platform used for profiling each sample.

We previously proposed a 7-gene signature predictive of SRS.^7,8^ We now asked whether this signature was applicable to RNA-seq by assessing the contribution of these genes to CC1 (**Figure 1B**). We observed significant contributions for 6 out of 7 genes (**Figure 1C**), demonstrating this signature is relevant to both microarray and RNA-seq. To assess if the signature can also be detected with rapid turn-around methods, we used quantitative reverse transcription polymerase chain reaction (qRT-PCR) to profile these genes in 115 samples from the GAinS study^7^, of which 72 and 94 also had microarray and RNA-seq measurements, respectively (**Methods** and **Supplementary Table 1**). We observed a very high level of agreement between profiling methods (**Figure 1D**), suggesting that the signature could potentially be used for qRT-PCR-based point-of-care testing.

An important limitation of our signature is its bias towards genes with higher expression in SRS2, with only one SRS1-associated gene (*TDRD9*). We reasoned that including more genes could make predictions more resilient. We combined our signature with a set of genes ranked amongst the top 1% with highest CC1 contribution (**Figure 1C** and **Methods**), which are expected to be robust to technological variation. This resulted in twelve additional genes, all of which showed comparable expression to the original signature (**Supplementary Figure 1A**).^7^ We henceforth refer to the 7-gene set as the *Davenport signature* and to the 19-gene set as the *Extended signature*.

We used these signatures to construct cross-platform reference maps of gene expression in sepsis. We compiled data from 1,044 patients in the GAinS study, profiled with up to three platforms, and integrated them based on our signatures. To make reference maps representative of a wider patient population, we also included three cohorts of healthy individuals (**Supplementary Table 1**). Integration was performed using mutual nearest neighbours (mNN), a method borrowed from single-cell omics which matches samples in one batch to their nearest neighbours in other batches (**Methods**).^16^ This resulted in two reference maps: the *Davenport map*, containing 3,264 samples (1,413 sepsis and 1,609 healthy), seven genes, and three modalities; and the *Extended map*, containing 3,149 samples (1,406 sepsis and 1,609 healthy), 19 genes, and two modalities. Samples in both reference maps clustered by endotype rather than technology, with the main axis of variation corresponding to a separation into healthy volunteer, SRS1, and SRS2 groups (**Figure 1E**). Thus, our reference maps capture a wide spectrum of transcriptional variation spanning health and critical illness.

### A classifier model for stratification of sepsis patients

We then used these signatures to predict endotypes. We split our reference maps into training (n=909) and test (n=2,355) sets, which we used to train random forest classifiers for each signature (**Methods**). Training sets contained all patient samples taken at ICU admission and for which SRS membership was known^7,8^ (n=639), as well as 270 randomly selected healthy volunteer samples, with all remaining samples allocated to the test set. Healthy volunteers were assigned to a newly defined SRS3 group. SRS3 is designed to capture healthy individuals, as well as patients in the low severity/recovery spectrum (i.e. transcriptionally closer to health).

Cross-validation revealed high accuracy across all endotype classes (AUROCs > 0.97; **Figure 2A**). This was recapitulated in the test set, with significant agreement between our predictions and previously proposed SRS labels for these samples.^7,8^ The Extended signature (Cohen’s Kappa = 0.91) marginally outperformed the Davenport signature (Cohen’s Kappa = 0.88). However, both reached a consensus for the majority of samples (97% and 84% agreement in microarray and RNA-seq, respectively; **Supplementary Figure 1B**). Predictions were consistent across technologies, with most samples assigned to the same SRS group regardless of profiling platform (**Figure 2B**). Thus, our models simultaneously achieve high accuracy and cross-technology applicability.

**Figure 2.**
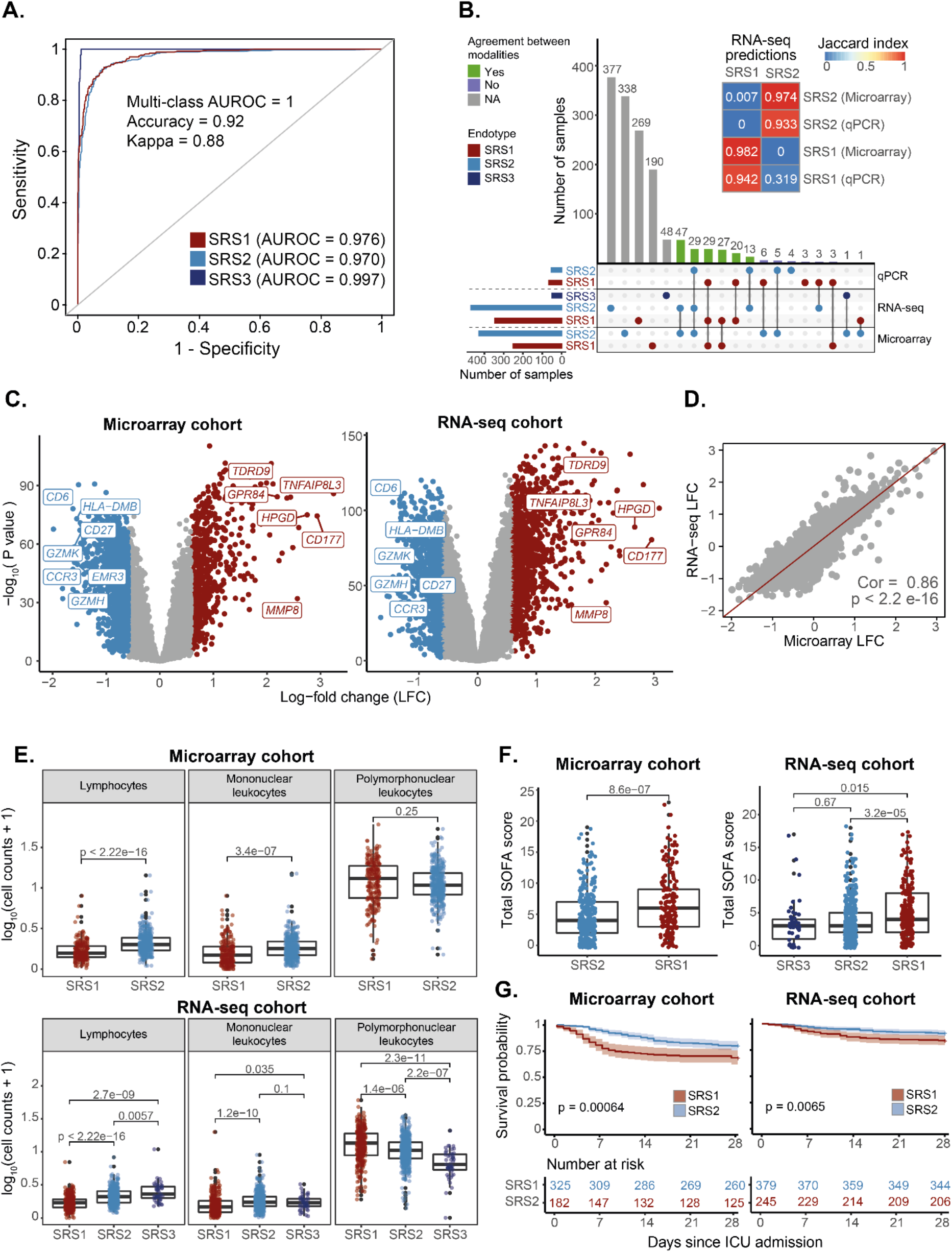
Stratification of sepsis patients in the GAinS cohort based on whole blood gene expression. **A)** Receiver operating characteristic (ROC) curves showing the performance of a random forest classifier in leave-one-out cross-validation. Lines of different colour indicate classification performance for different sepsis endotype classes. Areas under each ROC curve (AUROCs) are presented alongside an estimated multi-class AUROC. **B)** The UpSet plot (bottom) and heatmap (top) show the agreement between endotype labels derived from different gene expression profiling platforms. Bar heights in the UpSet plot indicate the number of samples assigned to each category, with dots and lines showing different types of overlap. Bar colours represent the predicted endotype class (horizontal bars) or whether there is agreement between labels derived from multiple modalities (vertical bars). Grey vertical bars indicate samples for which only one data modality was available. The colour scale in the heatmap indicates the overlap between two categories, as estimated using Jaccard indexes. **C)** Volcano plot showing differentially expressed genes between SRS1 and SRS2 groups. Each dot corresponds to a gene, for which its log-fold change (X axis) and log_10_ p value (Y axis) are indicated. Red dots represent genes significantly upregulated in the SRS1 endotype, with blue dots representing genes upregulated in the SRS2 group. Grey dots represent non-significant genes. Gene names were added to a subset of significantly differentially expressed genes of immune relevance. **D)** Correlation between log-fold changes between SRS1 and SRS2 estimated from microarray (X axis) and RNA-seq (Y axis) data. Each dot represents a gene. The identity line (red) is shown as a reference. Cor = Pearson correlation coefficient; p = correlation p value estimated using a two-tailed T test. **E)** Box plots comparing cell counts between SRS groups in the microarray (top) and RNA-seq (bottom) GAinS cohorts. Each dot represents a sample from a sepsis patient, colour coded by sepsis endotype. Each patient may contribute up to three samples, obtained at different times following ICU-admission. Box plots were defined in terms of medians (central line) and interquartile range (IQR; upper and lower box limits), with whiskers extending by ±1.5 the IQR from the limits of each box. p = p values from T tests (top) and Kruskal-Wallis tests (bottom). **F)** Box plots comparing SOFA scores between SRS groups in the microarray (left) and RNA-seq (right) cohorts. Dots represent samples obtained at the latest available time point from each patient, colour coded by endotype. Box plots are defined in terms of medians and IQR, with whiskers extending by ±1.5 the IQR. p = p values from either a T test (left) and a Kruskal-Wallis test (right). **G)** Kaplan-Meier curves comparing the 28-day survival of sepsis patients in the SRS1 and SRS2 groups. SRS groups were defined using samples from the latest available time point for each patient. Lines represent average survival probabilities (Y axis) of each endotype over time (X axis), with shaded areas indicating 95% confidence intervals. Patient numbers are shown at the bottom; p = p value from log-rank tests.

We next assessed if our models recapitulated known gene expression differences between endotypes. Differential expression analysis (**Methods**) revealed upregulation of neutrophil and innate immunity genes (e.g. *MMP8, GPR84*, and *CD177*) in SRS1, as well as downregulation of T cell function and antigen presentation genes (e.g. *CD27, CD6, CCR3*, and HLA class II molecules; **Figure 2C-D**). The top SRS1-associated pathways were Toll-like receptor (TLR) signalling, cytokine production, and glycolysis. In contrast, SRS2 was associated with T cell receptor (TCR) engagement, CD28-costimulation, and IFN*γ* signalling (**Supplementary Figure 2A**). This was supported by decreased lymphocyte and increased polymorphonuclear cell counts in SRS1 (**Figure 2E**). These observations agree with previous literature, demonstrating that our models distinguish biologically relevant molecular profiles.^7–9^

Finally, we asked whether SRS groups differed in clinical outcome. SRS1 patients showed higher Sequential Organ Failure Assessment (SOFA) scores, indicative of more severe organ dysfunction (**Figure 2F**). In the RNA-seq cohort, this was accompanied by increased Acute Physiology and Chronic Health Evaluation (APACHE) II scores (**Supplementary Figure 2B**). Survival analysis (**Methods**) further revealed that SRS1 patients are at increased risk of 28-day mortality (**Figure 2G**), demonstrating that our models can predict clinical outcomes.

### A quantitative score reflective of immune dysfunction

Sepsis can be viewed as a spectrum of illnesses with varying severities^17–19^, which raises the possibility of modelling patients as a continuum. We did this by using diffusion maps (**Methods**), a method designed to embed samples into a low-dimensional space that reflects their original connectivity.^20^ The first diffusion component (DC1) separated samples into a progression which started at SRS3 and gradually transitioned towards the SRS2 and SRS1 groups (**Figure 3A** and **Supplementary Figure 3A**). This provided evidence of the existence of a patient continuum.

**Figure 3.**
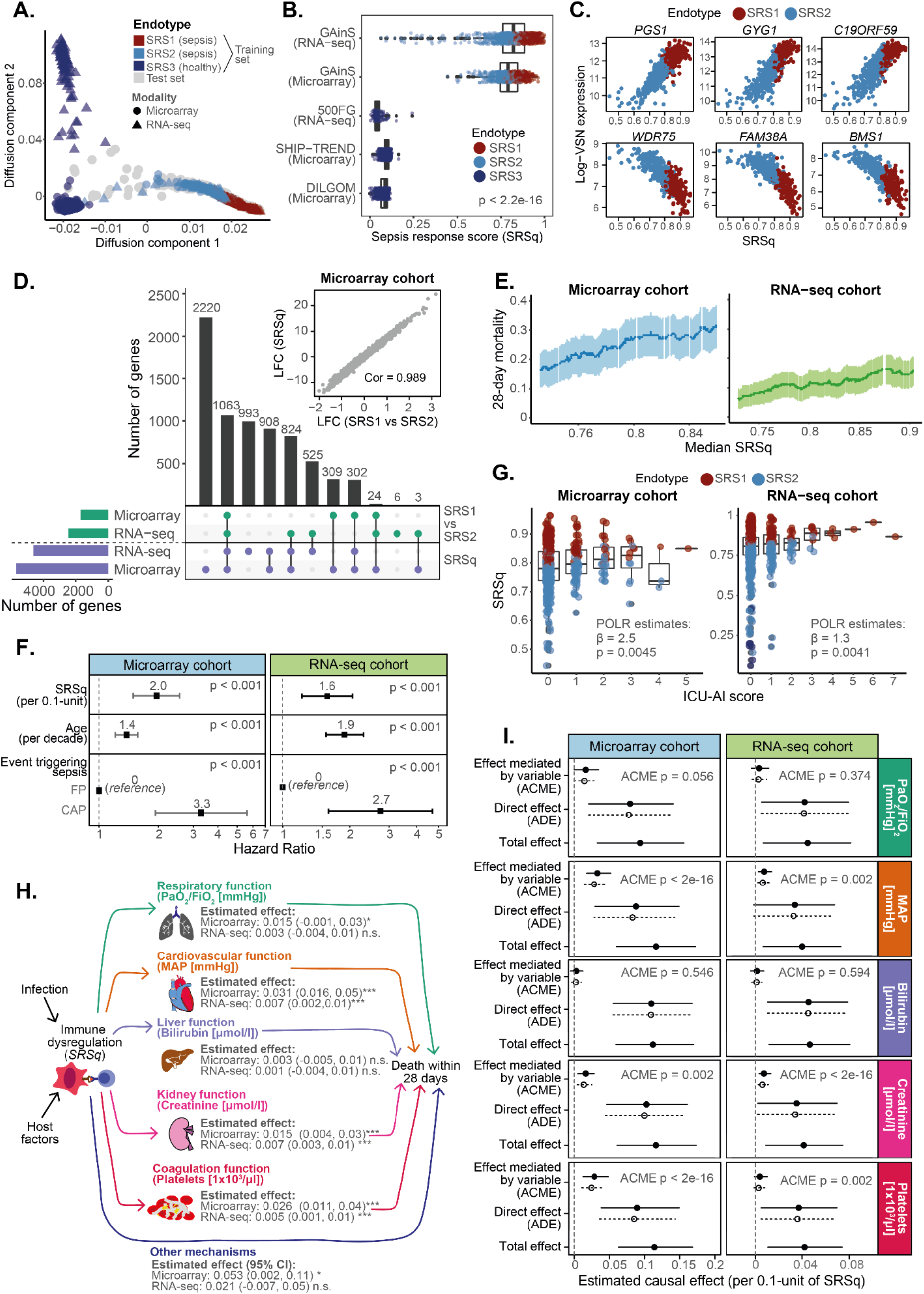
A quantitative score reflective of immune dysfunction and illness severity. **A)** Diffusion map plot showing the first two diffusion components based on the Extended reference map. Each dot represents a sample, with colours encoding endotype groups and shapes indicating the profiling platform used. **B)** Boxplots showing the distribution of quantitative sepsis response scores (SRSq) in each cohort within our reference map. SRSq was calculated by min-max scaling the first component of the diffusion map. Box plots are stratified by cohort. Each dot represents a sample, colour coded by sepsis endotype. Box plots were defined in terms of medians (central line) and interquartile ranges (IQR; upper and lower box limits), with whiskers extending by ±1.5 the IQR from the limits of each box. p = p value from a Kruskal-Wallis test. **C)** Top genes positively (top panels) and negatively (bottom panels) associated with SRSq. Each plot represents the expression level for the gene in question as measured using microarrays (log-VSN normalised intensities; Y axis) for different SRSq values (X axis). Each dot represents a sample, colour coded by sepsis endotype. **D)** Upset plot (bottom) and scatter plot (top) showing the agreement between differential gene expression along SRSq and differential gene expression between SRS1 and SRS2 endotypes. Bar heights in the UpSet plot indicate the number of genes called as differentially expressed in each comparison, with dots and lines showing different types of overlap. The scatter plot shows log-fold changes estimated between SRS1 and SRS2 (X axis) and along SRSq (Y axis) in microarray samples from the GAinS study. Each dot represents a gene; Cor = Pearson correlation coefficient. **E)** Association between SRSq and mortality in microarray (left) and RNA-seq (right) samples from GAinS. A sliding window was used to estimate the mortality of sets of samples with increasingly higher SRSq values. Dark dots represent 28-day mortality estimates and light lines show the associated 95% confidence intervals. **F)** Hazard ratio (HR) estimates obtained from applying a Cox Proportional Hazards model to microarray (left) and RNA-seq (right) samples in GAinS. Dots indicate the estimated HR for each variable, with lines showing 95% confidence intervals. **G)** Box plots showing the association between SRSq (Y axis) and ICU-acquired infection scores (ICU-AI; **Supplementary Table 2**) in microarray (left) and RNA-seq (right) samples from GAinS. Each dot represents a sample, colour coded by sepsis endotype. Box plots were defined in terms of medians (central line) and interquartile ranges (IQR; upper and lower box limits), with whiskers extending by ±1.5 the IQR from the limits of each box. ϐ = regression coefficient as estimated using proportional odds logistic regression (POLR); p = associated p value. **H)** Schematic diagram representing the structure of the causal model assumed for mediation analysis. Arrows represent hypothesised relationships, with arrow directions indicative of the direction of causality. Effect estimates from mediation analysis are provided for each arrow, along with their associated 95% confidence intervals and significance levels. Effect size estimates obtained from applying mediation analysis to microarray (left) and RNA-seq (right) samples in GAinS. Each row shows estimates obtained for a different clinical variable. Dots represent estimated effect sizes, with lines showing the associated 95% confidence interval. Solid and dotted lines represent effect sizes estimated with respect to samples with high SRSq (treatment condition) and low SRSq (control condition), respectively. ACME = Average Causal Mediation Effect; ADE = Average Direct Effect; p = mediation p value associated with ACME estimation.

We used DC1 to derive a quantitative metric reflective of the position of individuals along this continuum, which we refer to as the *quantitative sepsis response signature score* (SRSq; **Methods**). SRSq is bound between 0 and 1, with lower values indicating a patient is transcriptionally closer to health and higher values indicating similarity to the highest extreme of SRS1 (**Figure 3B**). SRSq scores derived using our two gene signatures were highly correlated (Pearson correlation = 0.84). However, the extended signature achieved better separation of healthy volunteers (**Supplementary Figure 3B**). To make SRSq calculation more straightforward, we trained machine learning models to predict this variable. We subdivided samples into training and test sets (as defined above) and trained random forest models for each gene signature (**Methods**). Model performance was high in both cross-validation and the test set (RMSE = 0.028; **Supplementary Figure 3D**), demonstrating reliable SRSq prediction.

We next investigated the molecular changes underlying SRSq. Both gene expression (**Figure 3C** and **Supplementary Figure 3D**) and cell counts (**Supplementary Figure 3E**) changed linearly as a function of SRSq, with lymphocyte counts decreasing proportionally to it. While both SRS and SRSq captured similar gene expression programs (99.7% of genes differentially expressed between SRS1 and SRS2 were also associated with SRSq; **Figure 3D**), our analysis identified 4,121 SRSq-associated genes which were not significantly different between SRS endotypes (**Figure 3C**). This more than doubled the associated gene set, demonstrating the power of modelling patients as a continuum.

We next investigated the relationship between SRSq and illness severity. We observed significant associations between SRSq and 28-day mortality (**Methods**) in both microarray and RNA-seq (**Figure 3E**), which were significant in Cox Proportional-Hazards models (**Methods**), even when accounting for age and source of sepsis (**Figure 3F**). Additionally, SRSq correlated with the severity of ICU-acquired infections (**Figure 3G**). Hazard ratios (HR = 2 and 1.6 in microarray and RNA-seq, respectively) indicated that a 0.1 increase in SRSq decreased patient survival as much as if the patient were a decade older. This illustrates the value of SRSq in risk estimation.

We subsequently investigated the processes linking immune dysfunction and death. Mediation analysis is a statistical method which takes candidate hypotheses and tests their compatibility with existing data by simulating how response variables change when other variables are altered one at a time.^21–23^ This is roughly equivalent to a computational randomised experiment. Results from mediation analysis suggest probable causal mechanisms, thus supporting hypothesis generation. We applied mediation analysis to SRSq and clinical measurements (**Methods** and **Supplementary Table 2**), assuming a model where SRSq influences organ dysfunction (i.e. SOFA scores), in turn increasing mortality (**Figure 3H**). This enabled us to estimate the direct effect of SRSq on death (i.e. how much mortality would increase if SRSq was raised while keeping SOFA constant), as well as its mediation effect (i.e. how much mortality would increase if SOFA was raised while keeping SRSq constant). Almost all the effect of SRSq on mortality was mediated by organ dysfunction (**Supplementary Figure 3F-G**).

We next assessed the role of individual organs by performing mediation analysis on all subcomponents of SOFA. The effect of SRSq on death was mediated by alterations in mean arterial pressure, coagulation (platelet counts), and, to a lesser extent, renal function (creatinine; **Figure 3H-I**). In contrast, we found no evidence of liver or lung dysfunction mediating this effect (**Figure 3I** and **Supplementary Figure 3H**), despite a large proportion of our cohort consisting of pneumonia patients. This suggests that systemic consequences of maladaptive inflammation in these patients relate more directly to SRSq than does lung damage.

### SepstratifieR: a machine learning framework for patient stratification

We next developed a software for SRS/SRSq prediction. We collected the models described above in an algorithmic framework called *SepstratifieR*. For a data set of interest, SepstratifieR: 1) extracts expression measurements for all signature genes (i.e. the Davenport or Extended signature, as specified by the user); 2) aligns samples to the corresponding reference map using mNN; and 3) predicts SRS and SRSq using random forest classifiers (**Figure 4**). This can be achieved in a single line of code, as described in https://github.com/jknightlab/SepstratifieR.

**Figure 4.**
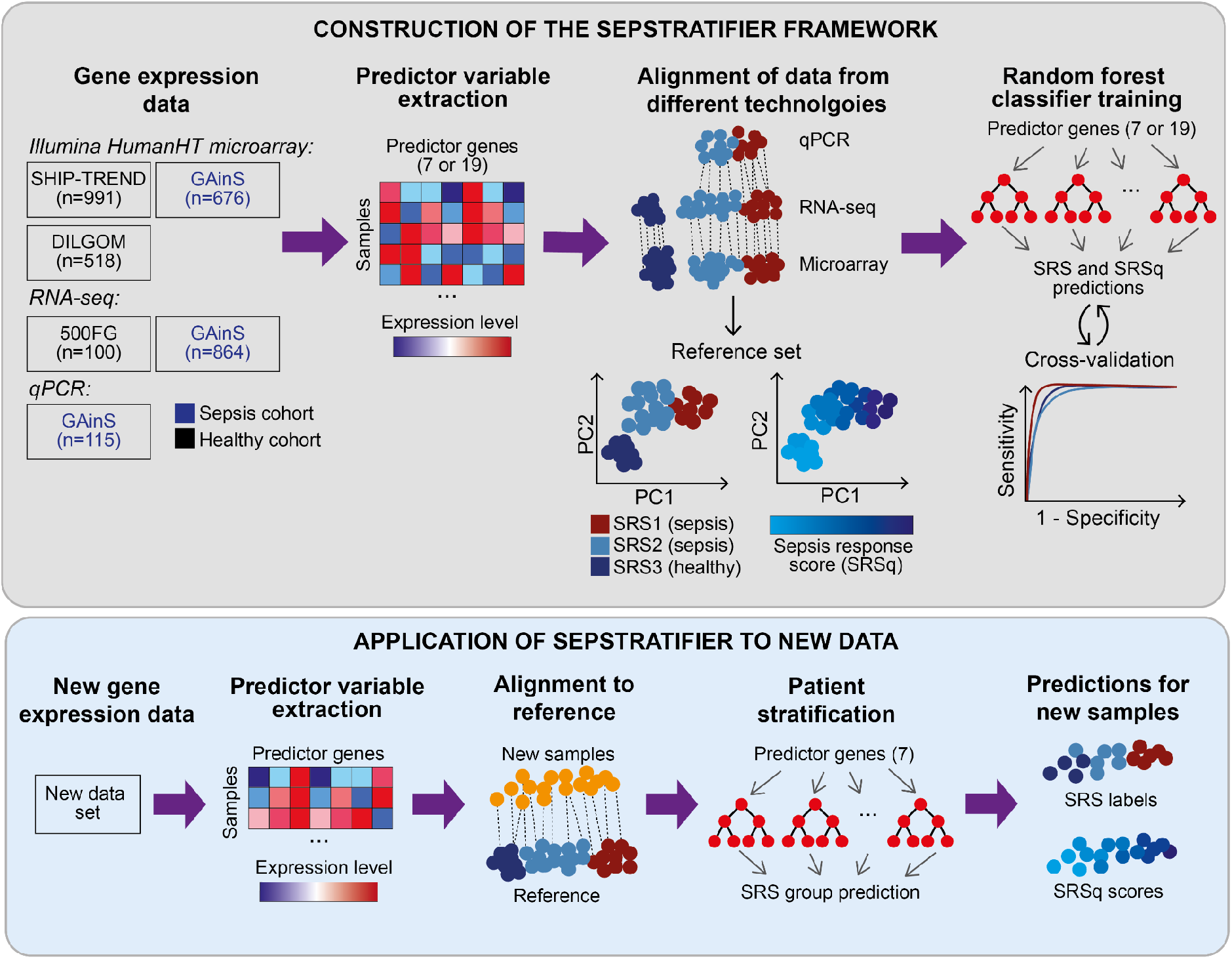
SepstratifieR’s construction and application to new data. Schematic representation of how the models within SepstratifieR were built (top panel) and how they are applied to new data (bottom panel). Publicly available data were used to construct sepsis reference maps based on small gene signatures by aligning different technologies into the same space. Next, random forest models were trained to predict SRS and SRSq, respectively. When applying SepstratifieR to a set of new samples, genes in the signature of interest are extracted and used to align the new samples to the sepsis reference map. After alignment, SRS and SRSq are predicted using pre-trained random forest models.

We evaluated the performance of SepstratifieR in two cohorts of sepsis patients (**Supplementary Table 1**). Exploratory analysis of a study by Parnell et al.^24^ revealed a clear separation between patients and controls, with sepsis patients segregating into survivor and non-survivor groups (**Supplementary Figure 4A**). These groups correlated with SepstratifieR’s predictions, with 77% of SRS2 patients surviving, compared to only 42% of SRS1 patients. Moreover, 82% of the SRS3 group consisted of healthy volunteers (**Supplementary Figure 4B**). These differences were also evident for SRSq, with elevated SRSq scores associated with higher APACHE II and mortality (**Supplementary Figure 4C and I**).

Gene expression differences agreed with previous observations, with all top SRSq-associated genes in GAinS recapitulated in this cohort and a high correlation of effect sizes observed between studies (**Supplementary Figure 4D-F**). Increased SRSq correlated with upregulation of innate immune pathways (e.g. inflammasome and IL-1 signalling), and glycolysis, as well as downregulation of TCR signalling, CD28-costimulation, and antigen presentation (**Supplementary Figure 4G**). This was supported by a positive correlation between SRSq and neutrophil proportions (**Supplementary Figure 4I**).

An important feature of this cohort was the availability of temporal information, with each patient profiled up to five times.^24^ This enabled us to model SRSq as a function of time and illness severity. While SRSq was constant in healthy individuals, it decreased over time in sepsis (**Supplementary Figure 4H**). This observation was driven by sepsis survivors (p value = 0.0032), with non survivors showing elevated SRSq scores throughout their disease trajectory (p value = 0.5). Thus, monitoring changes in SRSq over time might enable us to distinguish between different clinical trajectories.

We then applied SepstratifieR to the Molecular Diagnosis and Risk of Sepsis (MARS) study.^9^ MARS investigators previously described four endotypes, of which Mars1 showed higher mortality.^9^ Principal component analysis separated Mars1 patients along the first component (**Supplementary Figure 5**). In contrast, SRS groups predicted by SepstratifieR separated along the second component (**Supplementary Figure 5A**). Direct comparison revealed an overlap between SRS2 and Mars3, as well as an enrichment of Mars2 patients within SRS1. In contrast, 84% of the SRS3 group consisted of healthy volunteers (**Supplementary Figure 5B**). Surprisingly, the Mars1 endotype did not correspond to any of the groups identified by SepstratifieR, suggesting it represents an orthogonal axis of variation unrelated to our gene signatures. Finally, differential expression analysis showed highly correlated effect sizes between studies (Pearson correlation = 0.83) and a similar set of differentially active pathways (**Supplementary Figure 5C-E**).

At the clinical level, SRS1 patients in MARS showed a higher proportion of septic shock and elevated cardiovascular SOFA scores (**Supplementary Figure 5F-G**). However, we did not observe any differences in mortality between SRS groups (**Supplementary Figure 5H**) or along SRSq. This was surprising, given that SRS1 patients presented with higher rates of shock and organ dysfunction. We hypothesised that unmeasured factors could be severing the link between SRSq and death. For example, confounding or moderator variables could influence SRSq and mortality in opposite directions (**Supplementary Figure 5I**). We tested this using mediation analysis (**Methods**). While the total effect of SRSq on death was not significant, we observed significant mediation effects of SRSq on death via shock and SOFA scores (**Supplementary Figure 5J**). This suggests that an increase in SRSq leads to higher probabilities of shock and organ failure, which in turn increases mortality, but that unobserved variables might counterbalance this effect. For example, corticosteroid treatment has been shown to worsen the outcome of SRS2 patients who would otherwise be at lower risk.^12^

Thus, SepstratifieR can separate sepsis patients by molecular profile and predict illness severity in cohorts of different demographic and clinical composition.

### Stratification of H1N1 influenza patients by SRSq

SepstratifieR was trained on samples spanning a range of severities, from health to critical illness. This should extend its applicability to patients who do not fulfil sepsis criteria. To test this, we deployed SepstratifieR in a cohort of patients hospitalised with influenza.^25^ Exploratory analysis revealed a gradation of illness severities, with patients separating by degree of oxygen supplementation (**Figure 5A**). This gradation agreed with increases in SRSq scores (**Figure 5A**). At the molecular level, we confirmed increased expression of innate immunity genes proportionally to SRSq (**Figure 5B**), with a significant correlation of effect sizes between sepsis and influenza (Pearson correlation = 0.69; **Figure 5B-C**). This provided evidence that SRSq is applicable to acute viral infection.

**Figure 5.**
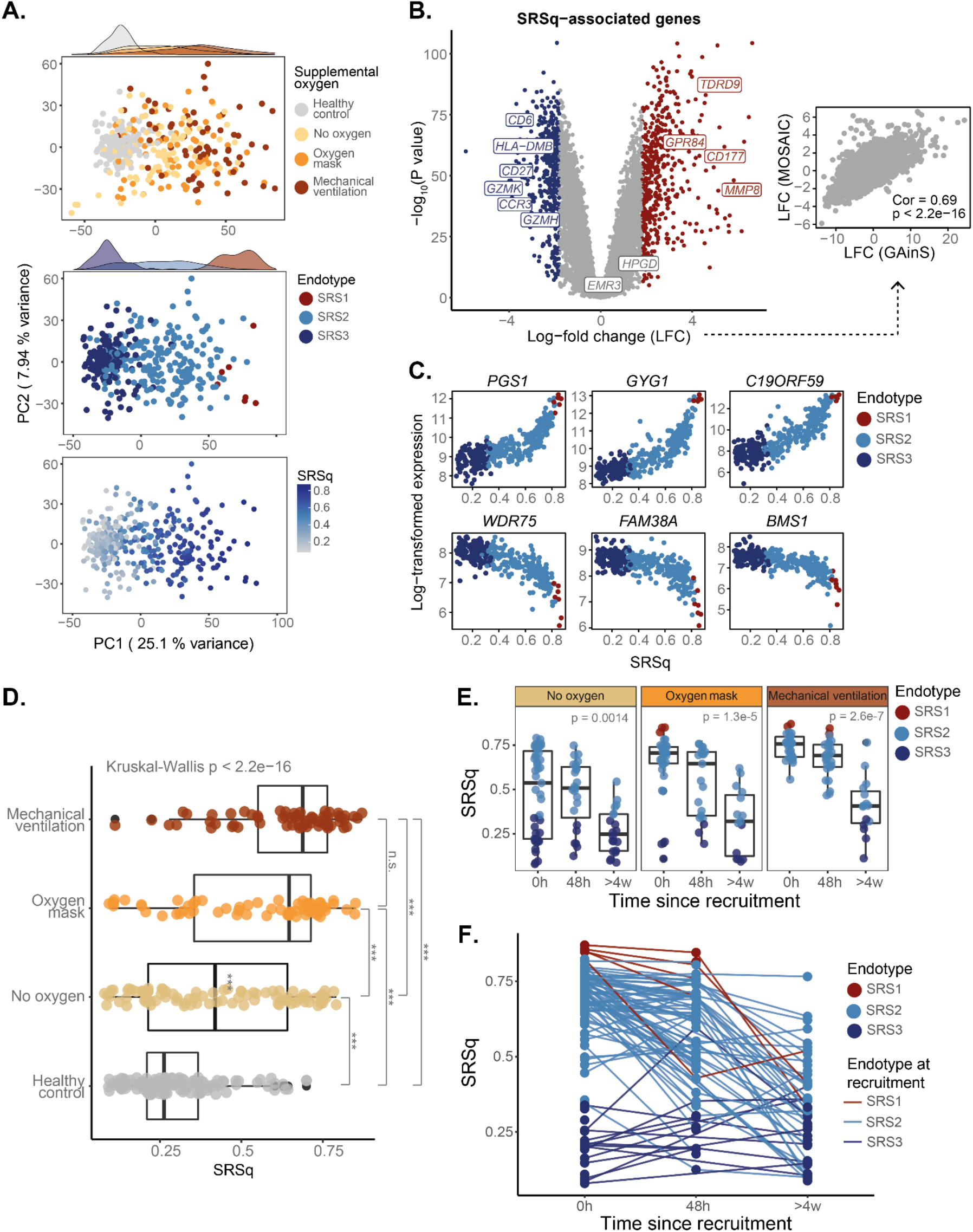
SRSq scores predict supplemental oxygen requirements and reveal temporal immune dynamics in H1N1 influenza. **A)** PCA plots based on whole blood transcriptomes. Each dot represents a sample, colour coded by: supplemental oxygen requirements (top panel), sepsis endotype (mid panel), and SRSq (bottom panel). Marginal distributions are provided where appropriate. **B)** Volcano plot (left) showing genes differentially expressed along SRSq. Each dot is a gene, with red indicating a positive and blue a negative association with SRSq. Grey dots represent genes which do not pass the significance threshold. Gene names are provided for a subset of significant genes with immune relevance. The associated scatter plot (right) shows the agreement between SRSq-associated log-fold changes in the GAinS study (X axis) and in Influenza patients (Y axis). Cor = Pearson correlation coefficient; p = correlation p value estimated using a two-tailed T test. **C)** Top genes positively (top panels) and negatively (bottom panels) associated with SRSq. Each plot represents the expression level for the gene in question (Y axis) for different SRSq values (X axis). Each dot represents a sample, colour coded by sepsis endotype. **D)** Box plots showing the association between SRSq (X axis) and supplemental oxygen requirement (Y axis). Each dot represents a sample. Box plots were defined in terms of median and IQR, with whiskers extending by ±1.5 the IQR from the limits of each box. Overall differences in SRSq distribution were tested using a Kruskal-Wallis test, followed by Dunn’s post-hoc test for each combination of categories (*** indicates a Bonferroni-adjusted Dunn’s test p < 0.01; n.s. Indicates a Bonferroni-adjusted Dunn’s test p > 0.05). **E)** Box plots showing the association between SRSq and time since patient admission. Each dot represents a sample, colour coded by sepsis endotype. Samples are stratified by oxygen requirement. Box plots are defined in terms of median and IQR, with whiskers extending by ±1.5 the IQR. p = p value from Kruskal-Wallis tests. **F)** Line plot showing changes of SRSq (Y axis) over time (X axis). Each dot represents a sample, colour coded by sepsis endotype. Lines join together samples from the same patient and are coloured based on the sepsis endotype assigned to that patient upon admission.

We next tested the association between SRSq and illness severity. We observed an increase in SRSq proportional to the extent of oxygen supplementation, with patients on mechanical ventilation showing an SRSq about 0.2 units higher than patients without supplemental oxygen (**Figure 5D**). In addition, we found a decrease of SRSq over time, with most patients displaying SRSq values equivalent to healthy volunteers after 4 weeks (**Figure 5E**). Interestingly, while patients with high SRSq upon admission (> 0.4) showed variable rates of SRSq decrease, patients with low initial SRSq (< 0.4) showed no changes over time (**Figure 5F**). These observations demonstrate that SepstratifieR is applicable to influenza, even when patients do not fulfil traditional sepsis criteria, and reveal complex patient trajectories.

### Stratification of COVID-19 patients by SRSq

Finally, we applied SepstratifieR to two COVID-19 cohorts: the COVID-19 Multi-Omic Blood Atlas (COMBAT)^26^ and the Deutsche COVID-19 Omics Initiative (DeCOI; **Supplementary Table 1**).^27^ In both instances, whole-transcriptome analysis showed a clear separation between patients and controls (**Figure 6A** and **Supplementary Figure 6A**). In COMBAT, patients further separated by severity (**Figure 6A**). While 90% of healthy volunteers in DeCOI were assigned to SRS3, 80% of COVID-19 samples were classified as either SRS1 or SRS2 (**Supplementary Figure 6B**). In COMBAT, the SRS3 group contained a mixture of healthy volunteers and community COVID-19 cases, who were never hospitalised. In contrast, SRS2 and SRS1 were enriched for patients with severe illness and in critical care, respectively (**Figure 6B**). SRSq increased proportionally to illness severity, with patients in critical care showing an SRSq 0.7 units higher than community cases (**Figure 6C**). This was recapitulated in DeCOI (**Supplementary Figure 6C**).

**Figure 6.**
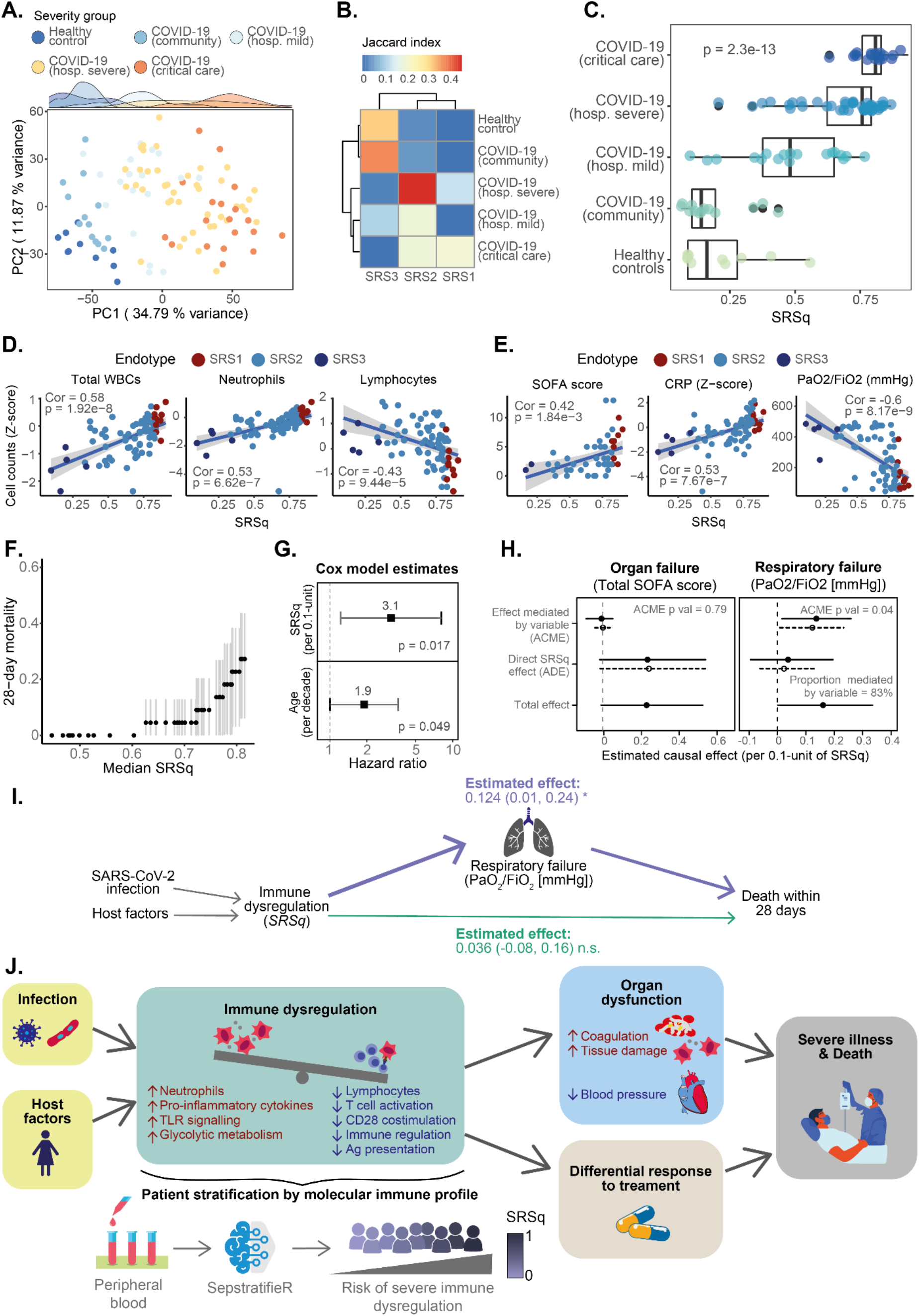
SRSq scores predict severity of illness and pinpoint mediators of morality in COVID-19. **A)** PCA plot based on whole blood transcriptomes. Each dot represents a sample, colour coded by its clinical severity group. Marginal distributions are provided. **B)** Heatmap showing the agreement between sepsis endotypes and clinical severity groups. Colours represent the level of agreement between two categories, as estimated using Jaccard indexes. **C)** Box plots showing the association between SRSq (X axis) and clinical severity (Y axis). Each dot represents a sample. Box plots were defined in terms of median and IQR, with whiskers extending by ±1.5 the IQR from the limits of each box. p = p value from a Kruskal-Wallis test. **D)** Association between SRSq (X axis) and cell counts (Z-scored; Y axis). Each dot represents a sample, coloured by sepsis endotype. Lines indicate the best linear fit, with shaded areas showing their associated 95% confidence intervals. Cor = Pearson correlation coefficient; p = correlation p value estimated using a two-tailed T test. **E)** Association between SRSq (X axis) and clinical variables (Y axis). Each dot represents a sample, coloured by sepsis endotype. Lines indicate the best linear fit, with shaded areas showing their associated 95% confidence intervals. Cor = Pearson correlation coefficient; p = correlation p value estimated using a two-tailed T test. **F)** Association between SRSq and mortality. A sliding window was used to estimate the mortality of sets of samples with increasingly higher SRSq values. Dark dots represent 28-day mortality estimates and light lines show the associated 95% confidence intervals. **G)** Hazard ratio (HR) estimates obtained from a Cox Proportional Hazards model. Dots indicate the estimated HR for each variable, with lines showing 95% confidence intervals. **H)** Effect size estimates obtained from mediation analysis, using total SOFA score (left) and P/F ratio (right) as mediator variables. Dots represent estimated effects, with lines showing their associated 95% confidence interval. Solid and dotted lines represent effect sizes estimated with respect to samples with high SRSq (treatment condition) and low SRSq (control condition), respectively. ACME = Average Causal Mediation Effect; ADE = Average Direct Effect; p = mediation p value associated with ACME estimation. **I)** Schematic diagram representing the causal model inferred from mediation analysis. Arrows represent causal relationships, with arrow directions indicative of the direction of causality. Effect estimates from mediation analysis are provided for each arrow, along with their 95% confidence intervals and significance levels. **J)** Graphical summary of SRSq and its biological interpretation.

We next compared transcriptional programs between sepsis and COVID-19. Differential expression analysis identified a similar set of SRSq-associated genes in both conditions (**Supplementary Figure 6D-E** and **Supplementary Figure 7A-B**). High SRSq scores in COVID-19 were associated with downregulation of antigen presentation, TCR signalling, and CD28-costimulation, as well as upregulation of TLR signalling, IL-1 signalling, and glycolysis (**Supplementary Figure 6F** and **Supplementary Figure 7C**). Furthermore, SRSq positively correlated with neutrophil and negatively correlated with lymphocyte counts (**Figure 6D** and **Supplementary Figure 6G**). Interestingly, elevated SRSq scores in COVID-19 were also associated with type I interferon signalling, a pathway not observed in sepsis.

We next evaluated the utility of SRSq for outcome prediction. SRSq was significantly associated with C reactive protein (CRP), respiratory function (P/F ratios), and SOFA scores, as well as with pneumonia indexes estimated by DeCOI investigators (**Figure 6E** and **Supplementary Figure 6H**). To assess if this resulted in differential outcomes, we evaluated the relationship between SRSq and 28-day mortality in COMBAT. While all patients with SRSq < 0.6 survived, we observed a sharp and linear increase in mortality in patients with SRSq > 0.6 (**Figure 6F**). This association was significant in a Cox Proportional-Hazards model (HR = 3.1 per 0.1-unit increase in SRSq), even when accounting for age (**Figure 6G**). Consequently, SRSq is predictive of clinical outcomes.

Finally, we performed mediation analysis assuming that immune dysfunction leads to higher rates of organ failure, in turn increasing mortality (**Figure 6H-I**). Surprisingly, we found no evidence of total SOFA scores mediating the effect of SRSq on death. Instead, this effect was entirely explained by respiratory function (i.e. P/F ratios; **Figure 6H-I**). While this analysis is limited by sample size, it suggests marked differences between causes of death in sepsis and COVID-19. In particular, in COMBAT, where SARS-CoV-2 infected patients span a wider severity range, respiratory failure plays a more prominent role, presumably because major systemic inflammation is not as prevalent.

In summary, SRSq is a quantitative score reflective of immune dysfunction and applicable to a variety of infections, including COVID-19. Elevated SRSq scores indicate decreased lymphocyte function and antigen presentation, increased neutrophil counts and TLR signalling, more severe illness, and higher likelihood of poor outcomes. This is likely explained by alterations in coagulation and blood pressure in sepsis, but by respiratory failure in COVID-19. These factors, possibly combined with a differential response to immunomodulatory therapy, increase patient mortality (**Figure 6J**).

## Discussion

We introduced SepstratifieR, a collection of models for stratification of patients with acute infection. SepstratifieR can be used in conjunction with numerous gene expression profiling methods and deployed across infections, successfully separating patients by severity and predicting clinical outcomes.

Our study addresses long-standing challenges. Firstly, it furthers our ability to identify endotypes at point of care. While this has been achieved in other diseases like inflammatory bowel disease^14,28^ and asthma^13^, similar methods are lacking for infectious disease. SepstratifieR provides a framework which can be used in conjunction with qRT-PCR testing, but remains applicable to full-transcriptome technologies. Secondly, SepstratifieR’s ability to model patients as a continuum makes it useful across a range of infections, independently of patients fulfilling sepsis criteria. This suggests that sepsis is one extreme along a spectrum of immune dysregulation. Interestingly, it was previously thought that systemic inflammatory response syndrome (SIRS), sepsis, and septic shock formed a continuum^17^, but evidence of different molecular processes operating in these conditions led to SIRS being removed as a diagnosis in Sepsis-3.^29,30^ While this simplifies diagnostic criteria, a spectrum of severities and mechanisms of immune dysfunction exists within sepsis^19^, and it is recognised that Sepsis-3 guidelines might fail to identify individuals with lower severity^18^. SRSq transcends diagnostic conventions and provides a quantitative metric of immune dysfunction, potentially supporting clinical decision making. Finally, SRSq describes molecular profiles better than endotypes, resulting in twice as many differentially expressed genes. Among the pathways relevant to SRSq is downregulation of antigen presentation, in agreement with immunosuppression being a hallmark of sepsis.^4^ This has also been observed in trauma, where suppressing antigen presentation might prevent responses to self-antigens after injury.^31,32^ Upregulation of TLR signalling was also associated with SRSq across all sources of infection. This agrees with life-threatening COVID-19 being associated with detrimental mutations in TLR and IFN genes^33,34^ and highlights the importance of this pathway in severe infection.

Nonetheless, it is worth highlighting some limitations. Firstly, SepstratifieR relies on bulk gene expression. Thus, it cannot establish which of the cellular alterations observed (e.g. decreased lymphocyte and increased neutrophil counts) cause immune dysfunction. We previously proposed T cell exhaustion as a key SRS1 feature.^7^ However, elevation of neutrophil extracellular traps in critical illness and detection of neutrophil signatures in severe COVID-19 suggest that neutrophils might also be dysregulated.^27,35,36^ Combining SRSq with single-cell technologies could help answer this question. Secondly, SRSq does not capture the full heterogeneity of sepsis. For example, some previously proposed endotypes (e.g. Mars1) do not correlate with SRSq.^9^ These orthogonal axes of variation could be lost when focusing on SRSq only. Finally, SepstratifieR relies on aligning new data to a reference set, which requires information from multiple samples. Thus, Sepstratifier cannot be applied to isolated samples, which currently limits its application in acute clinical settings. Further methodological developments are needed to extend this approach to individual samples while retaining its cross-technological scope.

In conclusion, SepstratifieR enables patient stratification in acute infection and models patients as a continuum. In combination with clinical biomarkers, SepstratifieR can enable precise risk estimation and inform the design and analysis of clinical trials, thus bringing us closer to precision approaches to treatment.

## Data Availability

All datasets used in this study are publicly available, with accession numbers listed in the manuscript.
All data produced in the present study are available upon request and will be made publicly available upon publication.

https://github.com/jknightlab/SepstratifieR

## Acknowledgements

This work was funded in whole, or in part, by the Medical Research Council (MR/V002503/1) (J.C.K. E.D.), Wellcome Trust Investigator Award (204969/Z/16/Z) (J.C.K), Wellcome Trust core funding to the Wellcome Sanger Institute (Grant number WT206194), Wellcome Trust Grants (090532/Z/09/Z and 203141/Z/16/Z) to core facilities Wellcome Centre for Human Genetics, and NIHR Oxford Biomedical Research Centre (J.C.K.). For the purpose of open access, the authors will apply a CC BY public copyright licence to any Author Accepted Manuscript version arising from this submission. We thank all the patients, patient families, nurses, and clinicians who participated in the GAinS study.

## Author contributions

JCK and EED designed the study. ECG performed data analysis. KLB, EED and JCK supervised the data analysis. DAS and HPS processed and provided clinical data. ECG, KLB, AK, DAS, DA, BPS, TVDP, ACG, CJH, EED and JCK discussed and interpreted the results. CG and ZHM performed qRT-PCR experiments and data processing. SM, CJH, JCK, and GAinS investigators performed patient recruitment. ECG, KLB, CG, ZHM, CJH, EED, and JCK wrote the manuscript.

## Methods

### Participants

#### The UK Genomics Advances in Sepsis (GAinS) cohort

Adult patients (≥18 yo) admitted into intensive care with sepsis due to community-acquired pneumonia (CAP, n=688) or faecal peritonitis (FP, n=358) were recruited through the UK Genomic Advances in Sepsis (GAinS) study (NCT00121196; https://ukccggains.com/) from 34 UK intensive care units (ICUs) between 16/11/2005 and 30/05/2018. Diagnoses were based on ACCP/SCCM guidelines.^37^ Inclusion and exclusion criteria are as described by Davenport et al.^7,8^

Ethics approval was granted nationally and locally, with informed consent obtained from all patients or their legal representative at the beginning of the study. This research was conducted under Research Ethics Committee approvals 05/MRE00/38, 08/H0505/78, and 06/Q1605/55.

### Procedures

#### Sample collection

Sample collection was performed as described by Davenport et al.^7,8^ Briefly, whole blood (∼10 mL) was obtained from patients on the first, third, and/or fifth day following ICU admission. Leukocyte isolation was performed at the bedside using the LeukoLOCK system (Thermo Scientific), with RNA extracted using the Total RNA Isolation Protocol (Ambion).

### Gene expression profiling

#### Microarray

Microarray-based profiling was performed on 676 samples (514 patients) from the GAinS study (**Supplementary Table 1**).^7,8^ Briefly, Illumina HumanHT-12 v4 expression BeadChips were used to quantify transcriptome-wide gene expression, with samples split into four batches.

Raw data were processed using GenomeStudio. Background subtraction, quality control, transformation, and normalisation were performed using the *vsn* package.^38^ This included identification of outliers and resolution of sample swaps based on genotyping data.^39^ Probes were filtered (detection p value < 0.05 in over 5% of samples) and measurements were averaged across all probes uniquely mapping to each gene. Batch effects were corrected using the empirical Bayes framework ComBat.^40^

#### RNA-sequencing

RNA-seq was performed on 864 samples (667 patients) from the GAinS study, including 134 samples with microarray data available (**Supplementary Table 1**). In brief, cDNA libraries were prepared using NEB Ultra II Library Prep kits (Illumina) and sequenced in a NovaSeq 6000 (Illumina). Reads were aligned to the reference genome (GRCh38 v99) using STAR (v2.7.3) and quantified using featureCounts.^41,42^ Counts were normalised and log-transformed, resulting in log-counts per million (log-CPM).

#### qRT-PCR

Seven genes predictive of SRS^7,8^ and two control housekeeping genes^43^ (*ACTB* and *TOP1*) were profiled using qRT-PCR in 115 RNA samples (107 patients) from the GAinS study (**Supplementary Table 1**).

Primers were designed using NCBI’s Primer-BLAST, ensuring that primers were exon-exon spanning, and amplicons were 70-250 bp and within 500 bp of a corresponding Illumina microarray probe. A Basic Local Alignment Search Tool (BLAST) search was performed against Ensembl GRCh37 v97 to ensure absence of homology to other genomic regions (E-value <0.01).

qRT-PCR was conducted on patient samples, two non-targeting controls (NTCs), and a healthy volunteer positive control, with cDNA generated from 500 ng of RNA using the LunaScript RT SuperMix Kit (New England Biolabs). Amplification was performed on 2 μL of cDNA in PCR reaction mix (2.5 µL of 10X PCR buffer, 0.75 µL of 50 mM MgCl_2_, 0.5 µL of 10 mM dNTP mix, 0.5 µL of 10 µM forward and reverse primers, 0.1 µL of PlatinumTM Taq DNA Polymerase, and nuclease-free water) using the following cycle: 1) 94°C for 2 min; 2) 40 cycles of 94°C for 30s, 55°C for 30s, and 72°C for 45s; and 3) 72°C for 5 min.

Raw data were processed with CFX Manager™ (BioRad) and C_q_ values <35 were removed. Mean C_q_ values for each gene were calculated by averaging over technical replicates, and normalisation to housekeeping genes was performed by subtracting the geometric mean of housekeeping genes from each gene in the SRS signature. Normalised C_q_ values were used for batch correction with *limma* (v3.44.1).^44^

#### Public data collection

Publicly available gene expression data were collected from three cohorts of healthy volunteers of German, Dutch and Finnish ancestry, as well as six infectious disease cohorts comprising all-cause sepsis, influenza and COVID-19 patients of different geographical origins (**Supplementary Table 1**).^9,24,25,27,45–50^ For microarray studies, probes were quality filtered (detection p value < 0.01 in over 20% of samples), and measurements were averaged across all probes uniquely mapping to each gene. For RNA-seq studies, counts were normalised and log-transformed and genes were quality filtered (>1 CPM in over 10% of samples).

#### Statistical analyses

#### Cross-technology data integration

Canonical correlation analysis (CCA) was performed using the sparse CCA algorithm in the *PMA* package (v1.2.1).^51^ Following CCA, datasets in the GAinS study were integrated with healthy volunteer data from three cohorts (**Supplementary Table 1**) based on a set of seven or 19 genes. Technical differences between cohorts were corrected using mutual nearest neighbours (mNN) with the *batchelor* package (v1.4.0).^16^ Batches were defined based on technology (i.e. Illumina HumanHT-12 arrays, RNA-seq, or qPCR).

### Construction of models for patient stratification

#### Definition of a quantitative sepsis response score

Diffusion maps were constructed using the *destiny* package.^52^ The first diffusion component (DC1) was used to derive a quantitative sepsis response score (SRSq), defined as the min-max scaled DC1 coordinate of each sample:

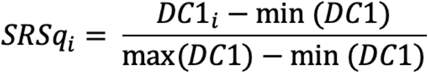

Where SRSq_i_ and DC1_i_ represent the sepsis response score and DC1 coordinates for the i-th sample, respectively. Min-max scaling restricted SRSq to the [0,1] range, where values closer to one indicate more severe immune dysfunction.

#### Random forest training and evaluation

Random forests were trained using Breiman’s algorithm with the *randomForest* (v4.6.14) and *caret* (v6.0.86) packages.^53,54,55^ Five hundred decision trees were built per forest, using either 7 or 19 genes as predictor variables. Either SRS labels or SRSq scores were used as response variables. Performance was evaluated using leave-one-out cross-validation (LOOCV) by estimating Cohen’s Kappa (for SRS) or root-mean-square errors (RMSE; for SRSq).^56^ The optimal number of predictor variables sampled at each split (i.e. ‘mtry’) was defined by assessing model performance over a range of values.

### Gene expression data analysis

#### Differential expression analysis

Differential gene expression between SRS endotypes and along SRSq was assessed using moderated T-tests with *limma*.^44^ Genes were called differential expressed whenever: 1) |fold-change| > 1.5 between SRS groups at a false discovery rate (FDR) of 0.05; or 2) |fold-change| > 3.5 per unit increase in SRSq at an FDR of 0.05 (i.e. a one-fold increase in expression for every 0.3 SRSq units).

#### Pathway enrichment analysis

Pathway enrichment was assessed using *XGR* (v1.1.8)^57^ using pathways listed in REACTOME.^58^ Pathways were deemed significantly enriched if they passed the 0.05 FDR threshold.

### Clinical data analysis

#### Pre-processing and integration with SRS/SRSq

GAinS clinical information was collected by local investigators using electronic case report forms (**Supplementary Table 2**). Data were quality filtered and assembled into a database for ease of access.

Associations between SRS and clinical variables were tested using Kruskal-Wallis one-way analysis of variance (for numeric variables) or Mood’s median test (for ordinal variables) with the *RVAideMemoire* package (v0.9.80).^59^ Associations between SRSq and clinical variables were assessed using correlation tests (for numeric variables) or proportional odds logistic regression (for ordinal variables) with the *MASS* package.^60,61^

#### Survival analysis

Mortality and time to death were censored (i.e. capped) at 28 days post ICU-admission. Kaplain-Meier curves were built using the *survival* (v3.1.12) package, modelling time to event as a function of SRS (with SRS measured at the latest time point available per patient). Visualisation was performed using *survminer* (v0.4.9)^62^, with significance estimated by log-rank tests.

To test for associations between SRSq and survival, samples were sorted by increasing SRSq (at the latest time point available) and a sliding window containing 35% of samples was used to estimate 28-day survival. Windows were slid one sample at a time using the *zoo* (v1.8.7) package^63^ until reaching the sample with highest SRSq. Survival within each window was estimated using *survival*. Associations were subsequently tested using Cox Proportional Hazard models.^64^ Hazard ratios (relative to 0.1-unit increases in SRSq) were modelled as an exponential function of SRSq, adjusting for age and source of sepsis (CAP or FP). Significance was assessed by log-rank tests.

#### Mediation analysis

Mediation analysis is best conceptualised using causal diagrams. In the diagram below, X represents an independent variable and Y a dependent variable, with M being a mediator lying in the causal path between X and Y. Arrows indicate the direction of causality:

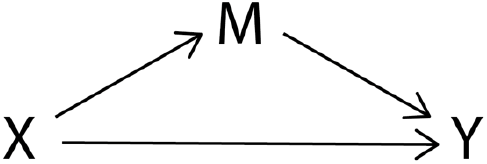

Mediation analysis estimates how much of the effect of X on Y depends on changes in M (the average causal mediation effect, ACME), as well as how much is independent of M (the direct causal effect, ADE).^21,65^ To do so, Y is modelled as a function of M and X, M itself being a function of X. ACME and ADE are then estimated either exactly or with simulations.^22,66^

In this study, SOFA scores, organ function measurements, and the presence/absence of shock were considered potential mediators of SRSq on mortality. Thus, each of them was modelled as a function of SRSq while accounting for covariates (age and source of sepsis). More specifically, numeric variables (e.g. SOFA scores) were modelled using a standard linear regression:

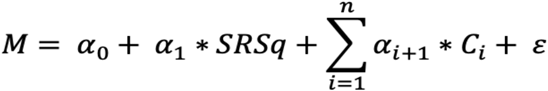

Where M is the mediator variable, Ci the i-th covariate, and *α* and *ε*the regression coefficients and random error term, respectively.

In contrast, binary variables (e.g. shock) were modelled using logistic regression with a probit model:

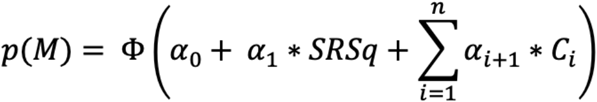

Where p(M) is the probability of the mediator, Ci the i-th covariate, *α* and *ε*the regression coefficients and random error term, respectively, and *Φ*the cumulative distribution function (CDF) of the normal distribution:

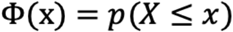

Assuming X is normally distributed with zero mean and unit standard deviation:

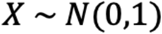

The dependent variable (i.e. mortality) was then modelled as a function of each mediator, one variable at a time, while accounting for SRSq and covariates. This was done using a logistic regression with probit model:

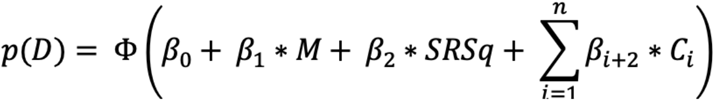

Where p(D) is the probability of death within 28 days of ICU admission, M the mediator of interest, Ci the i-th covariate, *α* and *ε*the regression coefficients and random error term, respectively, and *Φ*the cumulative distribution function (CDF) of the normal distribution.

ACMEs, ADEs, and their 95% confidence intervals were estimated using Monte Carlo simulations, as implemented in the *mediation* package (v4.5.0).^22,23,67^ Effect sizes were estimated relative to 0.1-unit increases in SRSq.

Mediation effects are best interpreted in terms of counterfactuals.^21^ In particular, ADEs tell us how much mortality would increase if SRSq were increased by 0.1 units while keeping the mediator constant. In contrast, ACMEs tell us how much mortality would increase if SRSq were held constant, but the mediator was artificially increased as if SRSq had increased by 0.1 units.

## Code and data availability

All codes will be made publicly available upon publication. The SepstratifieR package can be downloaded and installed from https://github.com/jknightlab/SepstratifieR.

Gene expression data for GAinS study samples are publicly available in ArrayExpress (E-MTAB-4421, E-MTAB-4451, E-MTAB-5273, and E-MTAB-5274). Accession numbers for all public datasets used are listed in **Supplementary Table 1**.

## Genomics Advances in Sepsis (GAinS) investigators

The following investigators, listed alphabetically by institution, were involved in patient recruitment, sample collection, and/or sample processing within the GAinS study:

Professor Nigel Webster, Dr Helen Galley, Jane Taylor, Sally Hall, Jenni Addison, Sian Roughton, and Heather Tennant (Aberdeen Royal Infirmary, Aberdeen, UK); Dr Achyut Guleri and Natalia Waddington (Blackpool Victoria Hospital, Blackpool, UK); Dr Dilshan Arawwawala, Dr John Durcan, Dr Alasdair Short, Karen Swan, Sarah Williams, Susan Smolen, and Christine Mitchell-Inwang (Broomfield Hospital, Chelmsford, UK); Dr Tony Gordon, Emily Errington, and Maie Templeton (Charing Cross Hospital, London); Dr Pyda Venatesh, Geraldine Ward, and Marie McCauley (Coventry and Warwickshire University Hospital, Coventry, UK); Dr Simon Baudouin and Charley Higham (Freeman Hospital, Newcastle, UK); Dr Jasmeet Soar, Sally Grier, and Elaine Hall (Frenchay Hospital, Bristol, UK and Southmead Hospital, Briston, UK); Dr Stephen Brett, David Kitson, Robert Wilson, Laura Mountford, and Juan Moreno (Hammersmith Hospital, London, UK); Dr Peter Hall and Jackie Hewlett (Huddersfield Royal Infirmary, Huddersfield, UK); Dr Stuart McKechnie, Dr Christopher Garrard, Dr Julian Millo, Dr Duncan Young, Paula Hutton, Penny Parsons, Alex Smiths, and Roser Faras-Arraya (John Radcliffe Hospital, Oxford, UK); Dr Jasmeet Soar and Parizade Raymode (Kettering General Hospital, Kettering, UK); Dr Jonathan Thompson, Sarah Bowrey, Sandra Kazembe, Natalie Rich, Prem Andreou, Dawn Hales, and Emma Roberts (Leicester Royal Infirmary, Leicester, UK); Dr Simon Fletcher, Melissa Rosbergen, and Georgina Glister (Norfolk and Norwich University Hospital, Norwich, UK); Dr Jeronimo Moreno Cuesta (North Middlesex Hospital, London, UK); Professor Julian Bion, Joanne Millar, Elsa Jane Perry, Heather Willis, Natalie Mitchell, Sebastian Ruel, Ronald Carrera, Jude Wilde, Annette Nilson, and Sarah Lees (Queen Elizabeth Hospital, Birmingham, UK); Dr Atul Kapila, Nicola Jacques, Jane Atkinson, Abby Brown, and Heather Prowse (Royal Berkshire Hospital, Reading, UK); Dr Anton Krige, Martin Bland, Lynne Bullock, and Donna Harrison (Royal Blackburn Hospital, Blackburn, UK); Dr Gary Mills, John Humphreys, and Kelsey Armitage (Royal Hallamshire and Northern General Hospitals, Sheffield, UK); Dr Shond Laha, Jacqueline Baldwin, Angela Walsh, and Nicola Doherty (Royal Preston Hospital, Preston, UK); Dr Stephen Drage, Laura Ortiz-Ruiz de Gordoa, and Sarah Lowes (Royal Sussex County Hospital, Brighton, UK); Dr Simon Baudouin, Charley Higham, Helen Walsh, Verity Calder, Catherine Swan, and Heather Payne (Royal Victoria Infirmary, Newcastle, UK); Dr David Higgins, Sarah Andrews, and Sarah Mappleback (Southend Hospital, Westcliff-on-Sea, UK); Professor Charles Hinds, Dr Chris Garrard, Dr D Watson, Eleanor McLees, and Alice Purdy (St Bartholomew’s Hospital and Royal London Hospital, London, UK); Dr Martin Stotz and Adaeze Ochelli-Okpue (St Mary’s Hospital, London, UK); Dr Stephen Bonner, Dr Iain Whitehead, Keith Hugil, Victoria Goodridge, and Louisa Cawthor (The James Cook University Hospital, Middlesbrough, UK); Dr Martin Kuper and Sheik Pahary (The Whittington Hospital, London, UK); Dr Geoffrey Bellingan, Dr Richard Marshall, Dr Hugh Montgomery, Jung Hyun Ryu, Georgia Bercades, and Susan Boluda (University College London Hospital, UCLH, London, UK); Dr Andrew Bentley, Katie Mccalman, and Fiona Jefferies (Wythenshawe Hospital, Manchester, UK); Professor Julian Knight, Dr Emma Davenport, Dr Katie Burnham, Dr Narelle Maugeri, Dr Jayachandran Radhakrishnan, Yuxin Mi, Alice Allcock, and Dr Cyndi Goh (Wellcome Centre for Human Genetics, University of Oxford, Oxford, UK).

## Supplementary figures

**Supplementary Figure 1.**
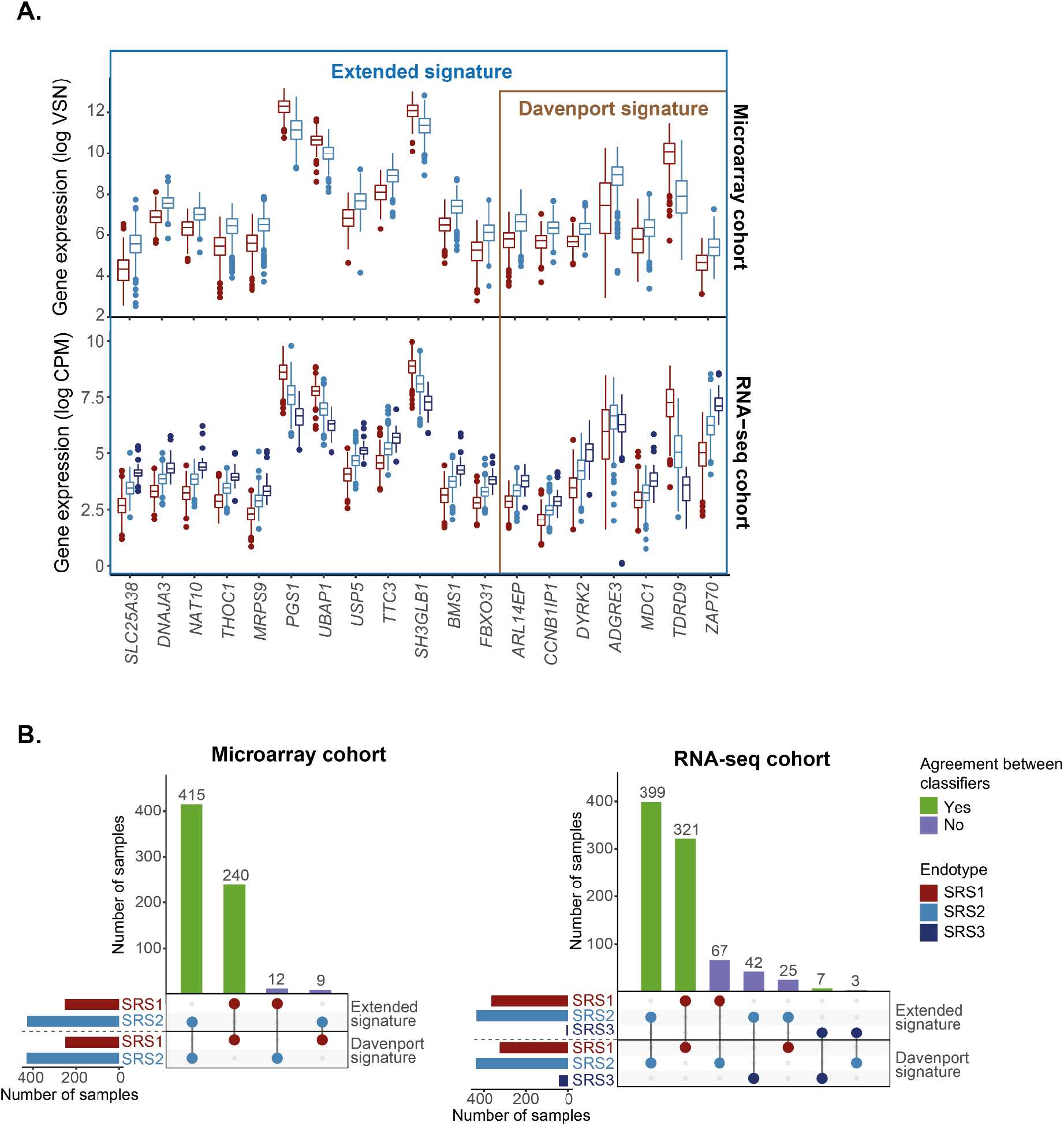
Two gene signatures for stratification of sepsis patients. **A)** Box plots showing the expression level (Y axis) of genes (X axis) in the Davenport and Extended signatures, as measured using microarrays (top panel) and RNA-seq (bottom panel). Each box plot summarises measurements from a group of patients, where patients are stratified by sepsis endotype (red = SRS1, light blue = SRS2, dark blue = SRS3). Box plots are defined in terms of medians (central line), IQR (box limits) and whiskers extending by ±1.5 the IQR. **B)** UpSet plot showing the agreement between endotype classifications obtained using the Davenport and Extended signature in microarray (left) and RNA-seq (right) samples of the GAinS study. Bar heights indicate the number of samples in each set, with dots and lines showing specific overlaps. Bar colours represent sepsis endotype groups (horizontal bars) and whether labels agree between both signatures (vertical bars).

**Supplementary Figure 2.**
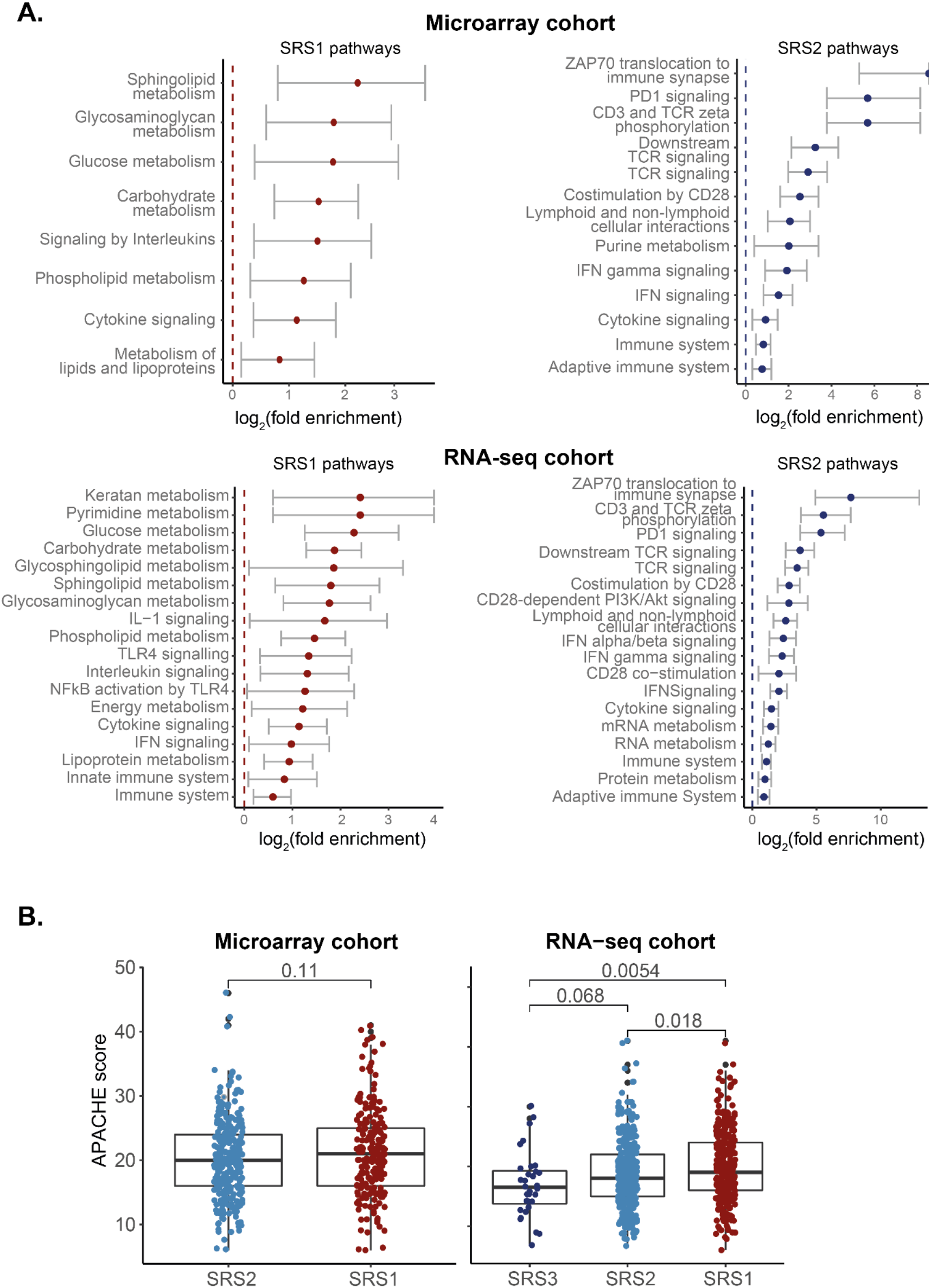
Features of sepsis response groups in the GAinS study. **A)** A subset of immune-relevant pathways significantly enriched for genes differentially expressed between SRS1 and SRS2 patients profiled using microarray (top panel) or RNA-seq (bottom panel). Each plot shows either pathways upregulated in the SRS1 (left) or SRS2 (right) endotype. Dots show the estimated log_2_-fold enrichment, with whiskers indicating the associated 95% confidence interval. All pathways were significant at FDR 0.05. **B)** Box plots showing difference in APACHE II scores between SRS endotypes in microarray (left) and RNA-seq (right) GAinS samples. Each dot represents a sample, colour coded by sepsis endotype. Box plots were defined in terms of medians (central line), IQR (box limits) and ±1.5 IQRs; p = p values from a T test (left) and a Kruskal-Wallis test (right).

**Supplementary Figure 3.**
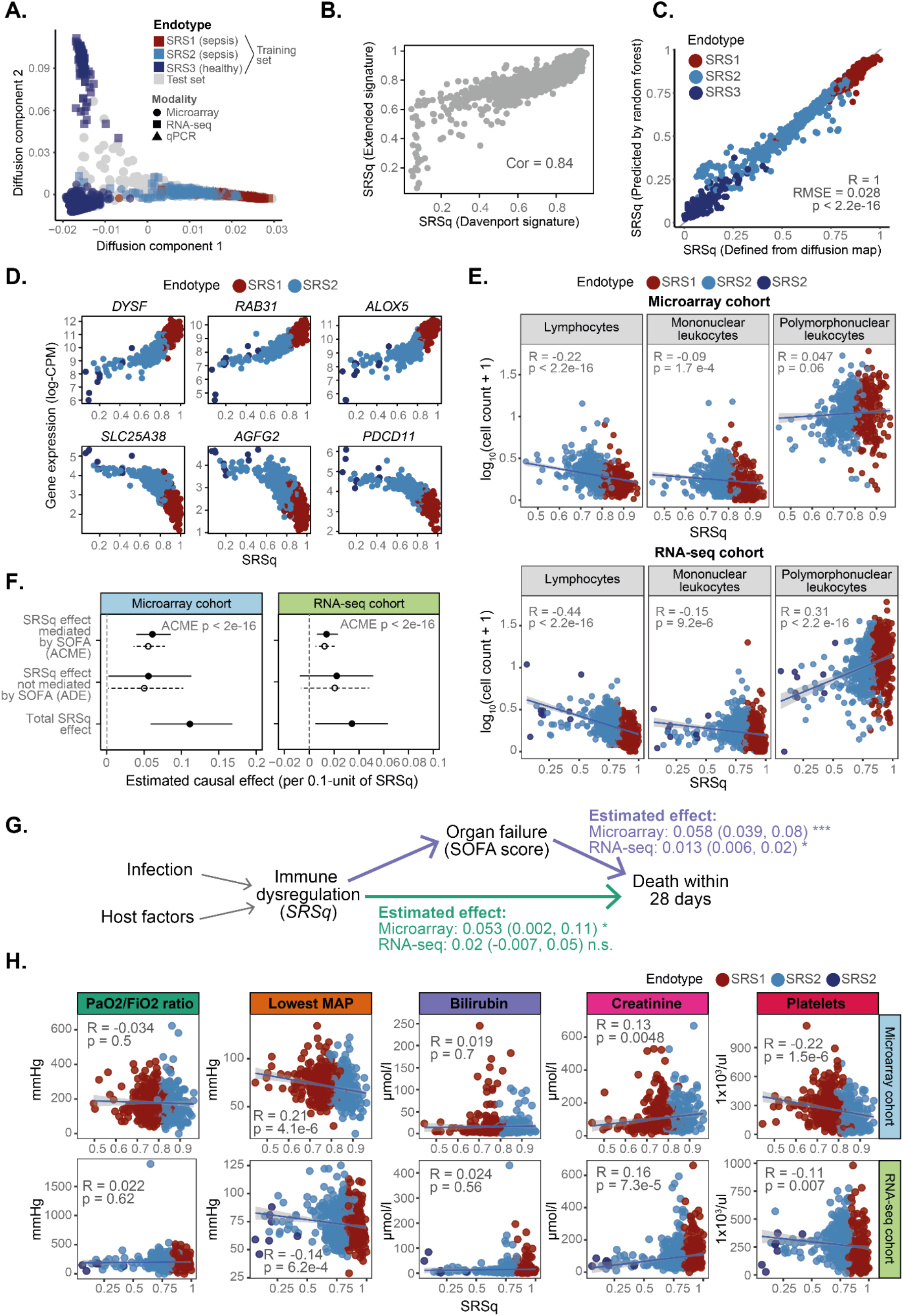
Clinical characteristics associated with SRSq in the GAinS study. **A)** Diffusion map plot showing the first two diffusion components based on the Davenport reference map. Each dot represents a sample, with colours encoding endotype groups and shapes indicating the profiling platform used. **B)** Correlation between SRSq values estimated using the Davenport (X axis) and Extended (Y axis) gene signatures. Each dot represents a sample. Cor = Pearson correlation coefficient. **C)** Correlation between SRSq scores as originally defined using diffusion maps (X axis) and SRSq scores as predicted with a random forest model (Y axis) based on the Extended gene signature. Each dot represents a sample, colour coded by sepsis endotype. R = Pearson correlation coefficient; RMSE = Root Mean Square Error; p = Correlation p value estimated using a two-tailed T test. **D)** Top genes positively (top panels) and negatively (bottom panels) associated with SRSq. Each plot represents the expression level for the gene in question as measured using RNA-seq (log-CPM; Y axis) for different SRSq values (X axis). Each dot represents a sample, colour coded by sepsis endotype. **E)** Association between SRSq (X axis) and log-transformed cell counts (Y axis)in GAinS microarray (top) and RNA-seq (bottom) samples. Each dot represents a sample, coloured by sepsis endotype. Lines indicate the best linear fit, with shaded areas showing their associated 95% confidence intervals. R = Pearson correlation coefficient; p = correlation p value estimated using a two-tailed T test. **F)** Effect size estimates obtained from applying mediation analysis to microarray (left) and RNA-seq (right) samples in GAinS, using SOFA score as a mediator variable. Dots represent estimated effect sizes, with lines showing the associated 95% confidence interval. Solid and dotted lines represent effect sizes estimated with respect to samples with high SRSq (treatment condition) and low SRSq (control condition), respectively. ACME = Average Causal Mediation Effect; ADE = Average Direct Effect; p = mediation p value associated with ACME estimation. **G)** Schematic diagram representing the causal model inferred from mediation analysis. Arrows represent causal relationships, with arrow directions indicative of the direction of causality. Effect estimates from mediation analysis are provided for each arrow, along with their 95% confidence intervals and significance levels. **H)** Correlation between SRSq (X axis) and clinical variables (Y axis) in GAinS microarray (top panels) and RNA-seq (bottom panel) samples. Rows show different clinical variables. Each dot represents a sample, colour coded by sepsis endotype. Lines indicate the best linear fit, with shaded areas showing the associated 95% confidence intervals. R = Pearson correlation coefficient; p = correlation p value estimated using a two-tailed T test.

**Supplementary Figure 4.**
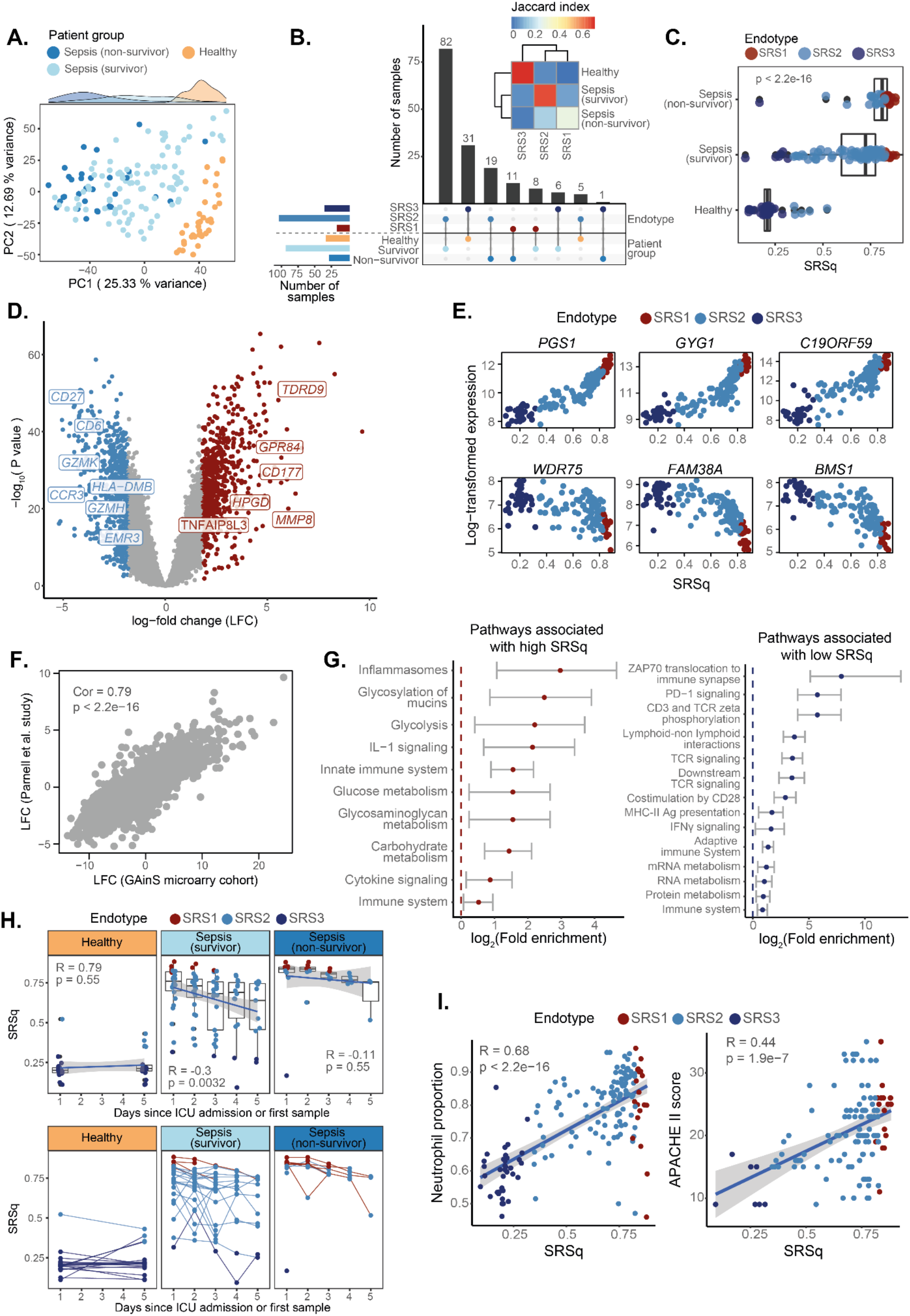
Validation of the utility of SRSq predictions in an independent cohort of sepsis patients. **A)** PCA plot based on whole blood transcriptomes. Each dot represents a sample, colour coded by its clinical group. Marginal distributions are provided. **B)** UpSet plot (bottom) and heatmap (top) showing the agreement between endotype labels and clinical groups. Bar heights in the UpSet plot indicate the number of samples assigned to each category, with dots and lines showing different types of overlap. Horizontal bar colours indicate endotype classes (top) and clinical groups (bottom). The colour scale in the heatmap shows the overlap between different endotypes and clinical groups as estimated using Jaccard indexes. **C)** Box plots showing the association between SRSq (X axis) and clinical group (Y axis). Each dot represents a sample. Box plots were defined in terms of median and IQR, with whiskers extending by ±1.5 the IQR; p = p value from a Kruskal-Wallis test. **D)** Volcano plot showing genes differentially expressed along SRSq. Each dot is a gene, with red indicating a positive and blue a negative association with SRSq. Grey dots represent genes which do not reach statistical significance. Gene names are provided for a subset of significant genes with immune relevance. **E)** Top genes identified as positively (top panels) and negatively (bottom panels) associated with SRSq in GAinS. Each plot represents the expression level for the gene in question in the study by Parnell et al. (Y axis) for different SRSq values (X axis). Each dot is a sample, colour coded by sepsis endotype. **F)** Scatter plot showing the correlation between SRSq-associated log-fold changes in the GAinS study (X axis) and in the study by Parnell et al. (Y axis); Cor = Pearson correlation coefficient; p = correlation p value estimated using a two-tailed T test. **G)** A subset of immune-relevant pathways significantly enriched for genes differentially expressed along SRSq. Plots show either pathways positively (left) or negatively (right) associated with SRSq. Dots show the estimated log_2_-fold enrichment, with whiskers indicating the associated 95% confidence interval. All pathways were significant at FDR 0.05. **H)** Correlation between SRSq and time of sampling, shown either as box plots (top panel) or line plots (bottom panel). In the box plots, each dot represents a sample colour coded by sepsis endotype. Box plots show average measurements for all samples corresponding to the same sampling time-point and are defined in terms of medians (central line), IQRs (box limits) and ±1.5 the IQR. Solid lines represent the best linear fit obtained when treating sampling time point as a numeric variable; R = Pearson correlation coefficient, p = correlation p values obtained using a two-tailed T test. In the line plots, each dot represents a sample colour coded by sepsis endotype, with lines connecting samples which correspond to the same patient repeatedly sampled over time. Lines are coloured based on the sepsis endotype assigned to each patient at the earliest time point available. **I)** Association between SRSq (X axis) and clinical variables (Y axis). Each dot represents a sample, coloured by sepsis endotype. Lines indicate the best linear fit, with shaded areas showing their associated 95% confidence intervals. R = Pearson correlation coefficient; p = correlation p value estimated using a two-tailed T test.

**Supplementary Figure 5.**
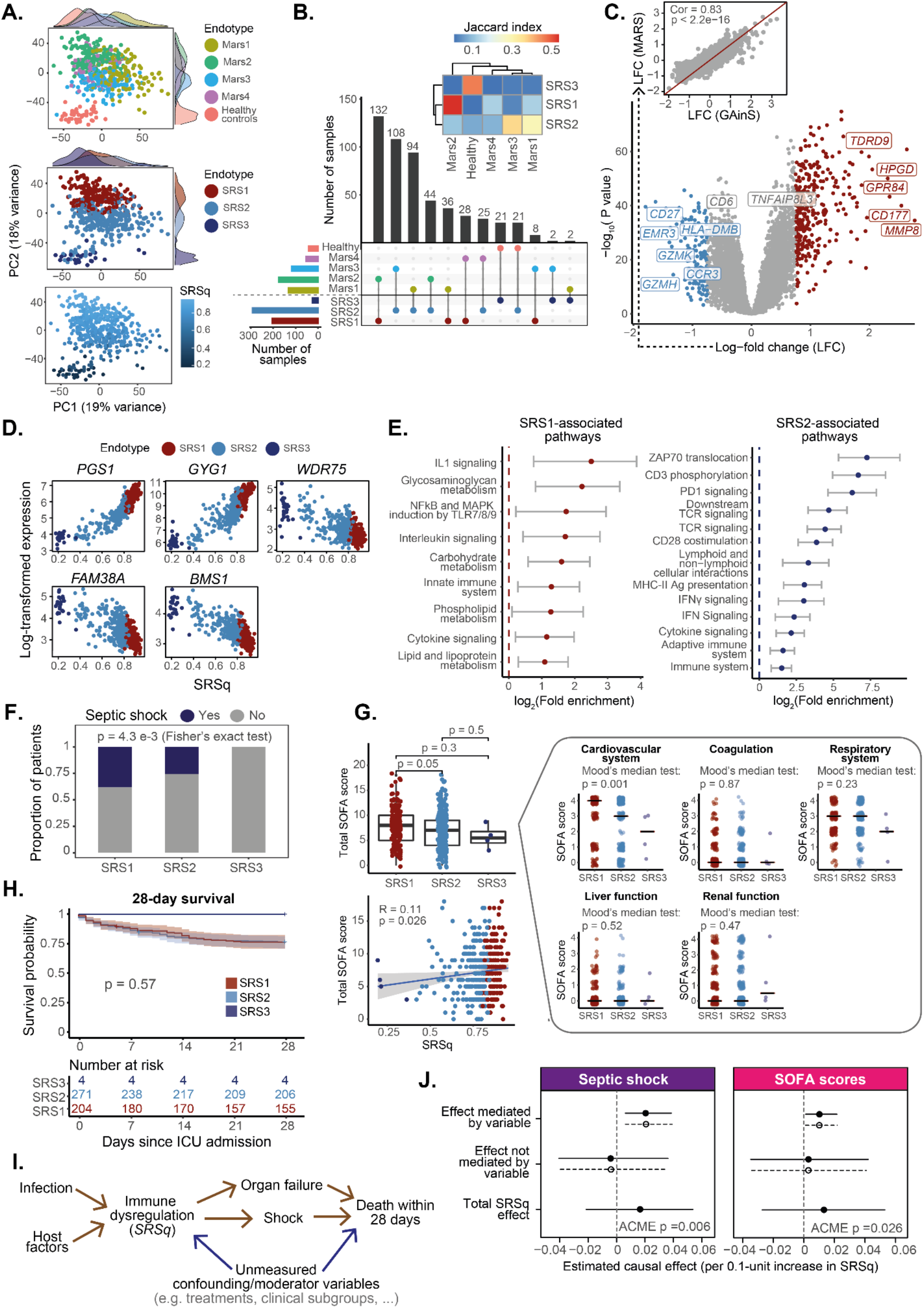
Prediction of SRSq scores in sepsis patients from the MARS study. **A)** PCA plots based on whole blood transcriptomes. Each dot represents a sample, colour coded by: Mars endotype (top panel), SRS endotype (mid panel), and SRSq (bottom panel). Marginal distributions are provided where appropriate. **B)** UpSet plot (bottom) and heatmap (top) showing the overlap between SRS and Mars endotype labels. Bar heights in the UpSet plot indicate the number of samples assigned to each category, with dots and lines showing different types of overlap. Horizontal bar colours indicate Mars (top) and SRS (bottom) endotype classifications, with dots and lines indicating specific overlaps. The colour scale in the heatmap shows the overlap between the two endotype definitions as estimated using Jaccard indexes. **C)** Volcano plot (bottom) showing genes differentially expressed between SRS1 and SRS2 samples. Each dot is a gene, with red indicating upregulation and blue downregulation in SRS1 samples. Grey dots show genes which do not reach statistical significance. Gene names are provided for a subset of significant genes with immune relevance. The associated scatter plot (top) shows the agreement between log-fold changes in the GAinS (X axis) and MARS (Y axis) studies. Cor = Pearson correlation coefficient; p = correlation p value estimated using a two-tailed T test. **D)** Top genes identified as positively (top panels) and negatively (bottom panels) associated with SRSq in GAinS. Each plot represents the expression level for the gene in question in the MARS cohort (Y axis) for different SRSq values (X axis). Each dot is a sample, colour coded by SRS endotype. **E)** A subset of immune-relevant pathways significantly enriched for genes differentially expressed between SRS1 and SRS2. Plots show either pathways upregulated (left) or downregulated (right) in SRS1 samples. Dots show the estimated log_2_-fold enrichment, with whiskers indicating the associated 95% confidence interval. All pathways were significant at FDR 0.05. **F)** Proportion of MARS patients diagnosed with septic shock in each SRS group. Bar heights indicate patient proportions per group, with colours corresponding to the presence or absence of shock; p = p value from Fisher’s exact test. **G)** Box plot showing the distribution of SOFA scores within each SRS group (top panel), and scatter plot showing the correlation between SOFA and SRSq scores (bottom panel). Each dot represents a sample, colour coded by SRS endotype. Box plots were defined in terms of medians (central line) and IQRs (box limits), with p values estimated using a Kruskal-Wallis test. The solid line in the scatter plot shows the best linear fit, with R = Pearson correlation coefficient and p = correlation p value obtained using a two-tailed T test. The associated plots show the distribution of organ-specific SOFA scores for each SRS group. Red lines indicate median values, with p values calculated using Mood’s median test. **H)** Kaplan-Meier curves comparing the 28-day survival of patients in the SRS1 and SRS2 groups. Each line represents average survival probabilities (Y axis) over time (X axis), with shaded areas indicating 95% confidence intervals. All patient numbers are shown at the bottom; p = p value from log-rank tests. **I)** Causal model used for mediation analysis. Arrows represent hypothesised relationships, with arrowheads indicative of the direction of causality. **J)** Effect size estimates obtained from mediation analysis, using either septic shock (left) or SOFA score (right) as the mediator variable. Dots represent estimated effect sizes, with lines showing the associated 95% confidence interval. Solid and dotted lines represent effect sizes estimated with respect to samples with high SRSq (treatment condition) and low SRSq (control condition), respectively. ACME = Average Causal Mediation Effect; ADE = Average Direct Effect; p = mediation p value associated with ACME estimation.

**Supplementary Figure 6.**
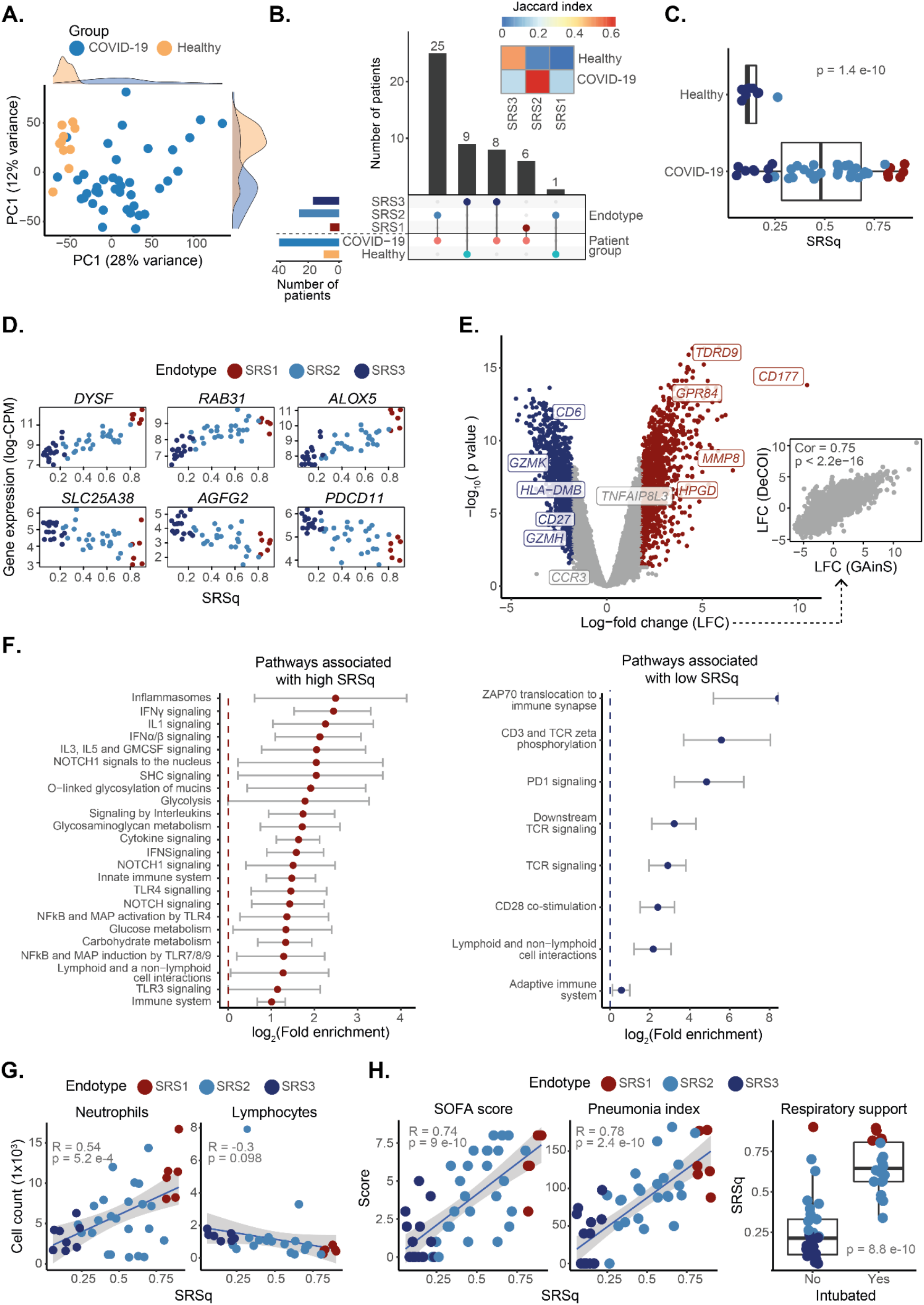
SRSq score prediction in COVID-19 patients from the DeCOI study. **A)** PCA plot based on whole blood transcriptomes. Each dot represents a sample, colour coded by its clinical group. Marginal distributions are provided. **B)** UpSet plot (bottom) and heatmap (top) showing the agreement between endotype labels and clinical groups. Bar heights in the UpSet plot indicate the number of samples assigned to each category, with dots and lines showing different types of overlap. Horizontal bar colours indicate endotype classes (top) and clinical groups (bottom). The colour scale in the heatmap shows the overlap between endotypes and clinical groups as estimated using Jaccard indexes. **C)** Box plots showing the association between SRSq (X axis) and clinical group (Y axis). Each dot represents a sample, colour coded by sepsis endotype (red = SRS1, light blue = SRS2, dark blue = SRS3). Box plots were defined in terms of median and IQR, with whiskers extending by ±1.5 the IQR; p = p value from a Kruskal-Wallis test. **D)** Top genes identified as positively (top panels) and negatively (bottom panels) associated with SRSq in GAinS. Each plot represents the expression level for the gene in question in the DeCOI study (Y axis) for different SRSq values (X axis). Each dot is a sample, colour coded by sepsis endotype. **E)** Volcano plot showing genes differentially expressed along SRSq. Each dot is a gene, with red indicating a positive and blue a negative association with SRSq. Grey dots represent genes which do not reach statistical significance. Gene names are provided for a subset of significant genes with immune relevance. The associated scatter plot (right) shows the agreement between log-fold changes in the GAinS (X axis) and DeCOI (Y axis) studies. Cor = Pearson correlation coefficient; p = correlation p value estimated using a two-tailed T test. **F)** A subset of immune-relevant pathways significantly enriched for genes differentially expressed along SRSq. Plots show either pathways positively (left) or negatively (right) associated with SRSq. Dots show the estimated log_2_-fold enrichment, with whiskers indicating the associated 95% confidence interval. All pathways were significant at FDR 0.05. **G)** Association between SRSq (X axis) and cell counts (Z-scored; Y axis). Each dot represents a sample, coloured by sepsis endotype. Lines indicate the best linear fit, with shaded areas showing their associated 95% confidence intervals. Cor = Pearson correlation coefficient; p = correlation p value estimated using a two-tailed T test. **H)** Association between SRSq (X axis) and clinical variables (Y axis). Each dot represents a sample, coloured by sepsis endotype. Lines indicate the best linear fit, with shaded areas showing their associated 95% confidence intervals, with Cor = Pearson correlation coefficient and p = correlation p value estimated using a two-tailed T test. Box plots were defined in terms of median and IQR, with whiskers extending by ±1.5 the IQR and p = p value from a T test.

**Supplementary Figure 7.**
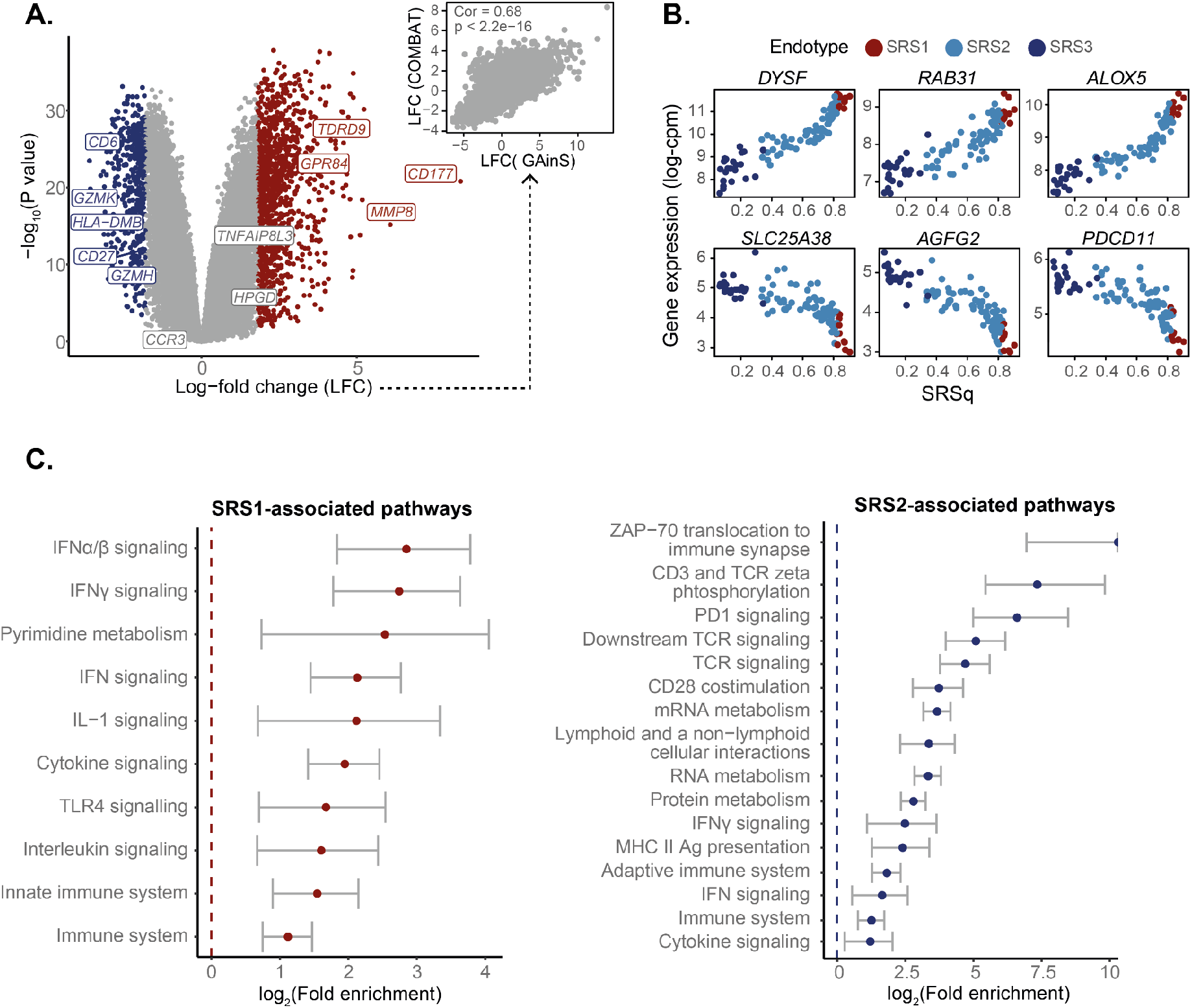
Transcriptional profiles associated with SRSq in COVID-19 patients from the COMBAT study. **A)** Volcano plot showing genes differentially expressed along SRSq. Each dot is a gene, with red indicating a positive and blue a negative association with SRSq. Grey dots represent genes which do not reach statistical significance. Gene names are provided for a subset of significant genes with immune relevance. The associated scatter plot (right) shows the agreement between log-fold changes in the GAinS (X axis) and COMBAT (Y axis) studies. Cor = Pearson correlation coefficient; p = correlation p value estimated using a two-tailed T test. **B)** Top genes identified as positively (top panels) and negatively (bottom panels) associated with SRSq in GAinS. Each plot represents the expression level of the gene in question in the COMBAT study (Y axis) for different SRSq values (X axis). Each dot is a sample, colour coded by sepsis endotype. **C)** A subset of immune-relevant pathways significantly enriched for genes differentially between sepsis endotypes. Plots show either pathways upregulated (left) or downregulated (right) in SRS1 samples. Dots show the estimated log_2_-fold enrichment, with whiskers indicating the associated 95% confidence interval. All pathways were significant at FDR 0.05.

## Supplementary tables

**Supplementary Table 1.**
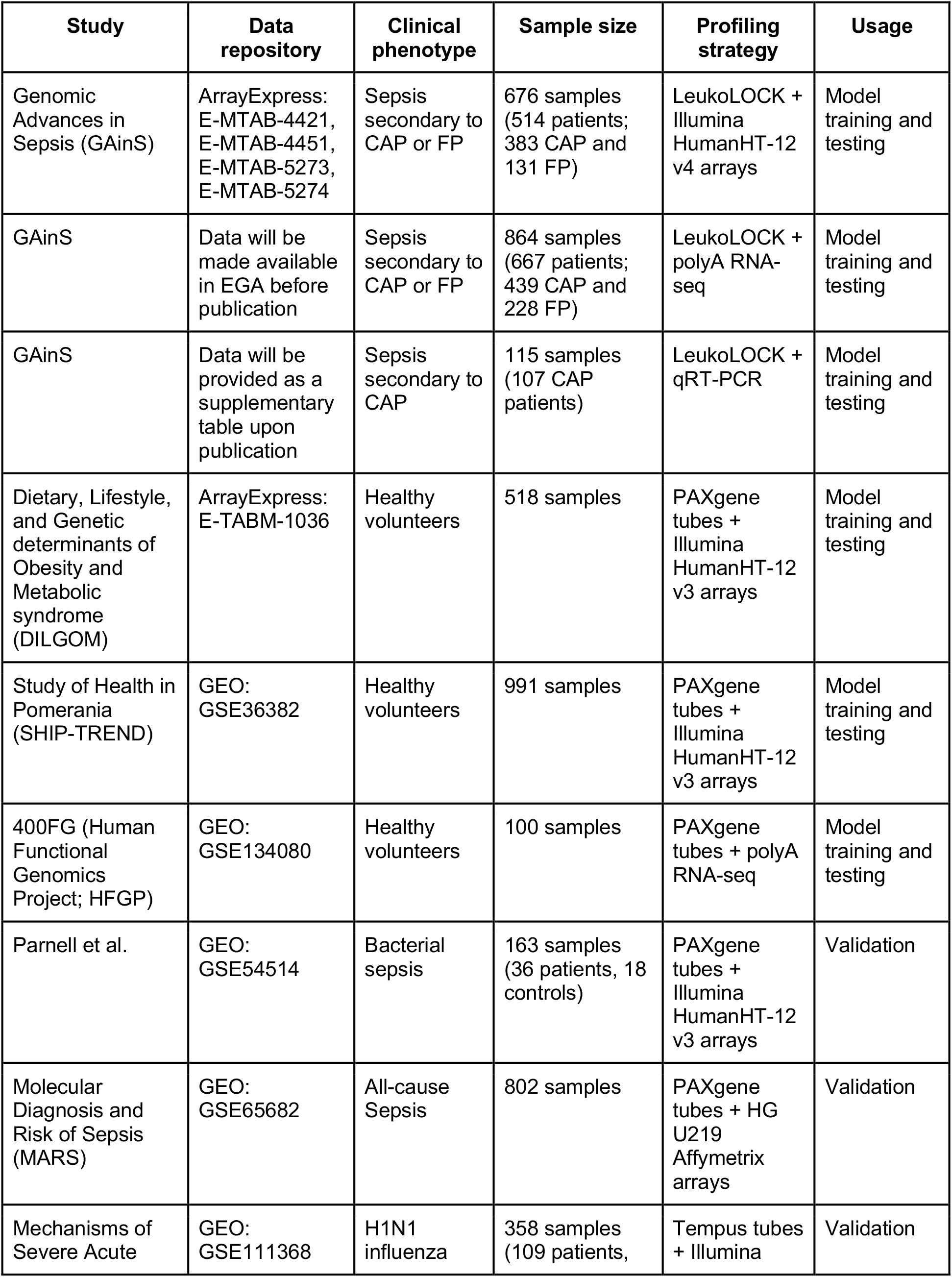
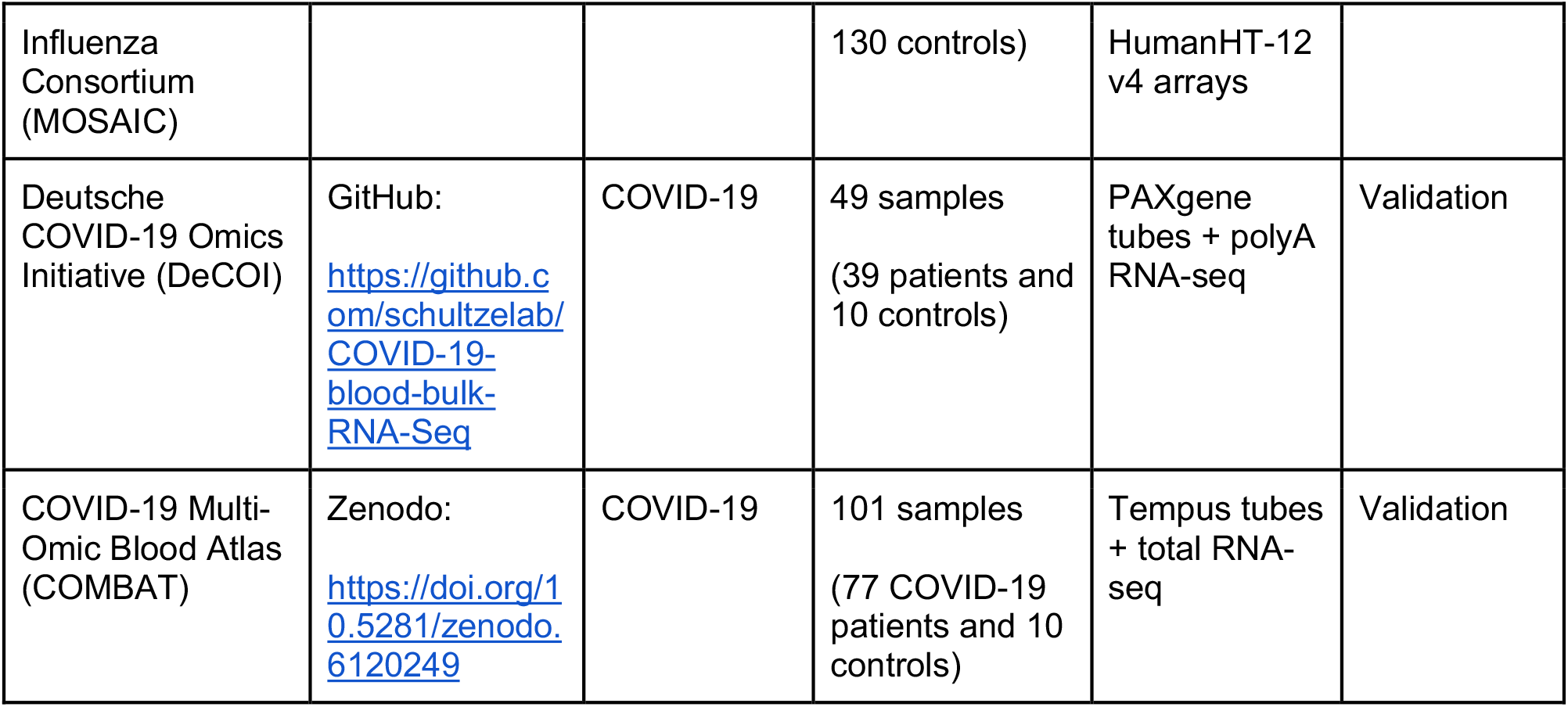
Publicly available data sets used for model training and validation. List of all publicly available gene expression data sets used throughout this study, either for model training or for validation of results. All data sets consisted of whole blood transcriptomes.

**Supplementary Table 2.** Clinical variables in the GAinS study. List of clinical and outcome variables available for the GAinS cohort and used throughout this study. Different columns specify how these variables were measured and in which units.

| Clinical variable | Definition | Variable type | Unit |
| --- | --- | --- | --- |
| Mortality event | Information on whether the patient died (1) or survived (0) | Binary | Yes/No |
| Time to mortality event | Number of days from ICU admission to death (if it occurred) | Numeric (integer) | Days |
| Cell counts | Full blood counts as measured in hospital using haematology analysers | Numeric (float) | $1 \times 10^9$ cells/l |
| APACHE scores | Total Acute Physiology and Chronic Health Evaluation (APACHE) II scores measured at ICU admission | Numeric (integer) | 0-24 points |
| Total SOFA scores | Total Sequential Organ Failure Assessment (SOFA) scores measured during the first, third and/or fifth days after ICU admission | Numeric (integer) | 0-71 points |
| Mean arterial pressure (MAP) | Lowest mean arterial pressure measurement obtained for each patient | Numeric (float) | mmHg |
| PF ratio | $\text{PaO}_2/\text{FiO}_2$ ratio (i.e. Horowitz index) | Numeric (float) | mmHg |
| Bilirubin levels | Highest bilirubin measurement obtained for each patient | Numeric (float) | $\mu\text{mol/l}$ |
| Creatinine levels | Highest creatinine measurement obtained for each patient | Numeric (float) | $\mu\text{mol/l}$ |
| Platelet levels | Number of platelets measured | Numeric (float) | $1 \times 10^3$ platelets/$\mu\text{l}$ |
| ICU-AI scores | ICU-acquired infection score.<br>This score reflects how many of the following ICU-acquired complications were diagnosed for each patient: 1) ventilator associated pneumonia (VAP), 2) lower respiratory tract infection, 3) bacteraemia, 4) line-related infection, 5) wound infection, 6) urinary tract infection (UTI), 7) other infection | Ordinal (integer) | 0-7 points |

## References

1. Vos, T. et al. Global burden of 369 diseases and injuries in 204 countries and territories, 1990–2019: a systematic analysis for the Global Burden of Disease Study 2019. Lancet 396, 1204–1222 (2020).

2. Chowell, G. et al. Severe respiratory disease concurrent with the circulation of H1N1 influenza. N. Engl. J. Med. 361, 674–679 (2009).

3. Gallo Marin, B. et al. Predictors of COVID-19 severity: A literature review. Rev. Med. Virol. 31, 1–10 (2021).

4. van der Poll, T., van de Veerdonk, F. L., Scicluna, B. P. & Netea, M. G. The immunopathology of sepsis and potential therapeutic targets. Nat. Rev. Immunol. 17, 407–420 (2017).

5. Rudd, K. E. et al. Global, regional, and national sepsis incidence and mortality, 1990-2017: analysis for the Global Burden of Disease Study. Lancet 395, 200–211 (2020).

6. Sandhu, C., Qureshi, A. & Emili, A. Panomics for Precision Medicine. Trends Mol. Med. 24, 85–101 (2018).

7. Davenport, E. E. et al. Genomic landscape of the individual host response and outcomes in sepsis: a prospective cohort study. Lancet Respir Med 4, 259–271 (2016).

8. Burnham, K. L. et al. Shared and Distinct Aspects of the Sepsis Transcriptomic Response to Fecal Peritonitis and Pneumonia. Am. J. Respir. Crit. Care Med. 196, 328– 339 (2017).

9. Scicluna, B. P. et al. Classification of patients with sepsis according to blood genomic endotype: a prospective cohort study. Lancet Respir Med 5, 816–826 (2017).

10. Sweeney, T. E. et al. A community approach to mortality prediction in sepsis via gene expression analysis. Nat. Commun. 9, 694 (2018).

11. Baghela, A. et al. Predicting sepsis severity at first clinical presentation: The role of endotypes and mechanistic signatures. EBioMedicine 103776 (2022).

12. Antcliffe, D. B. et al. Transcriptomic Signatures in Sepsis and a Differential Response to Steroids. From the VANISH Randomized Trial. Am. J. Respir. Crit. Care Med. 199, 980– 986 (2019).

13. Bhakta, N. R. et al. A qPCR-based metric of Th2 airway inflammation in asthma. Clin. Transl. Allergy 3, 24 (2013).

14. Biasci, D. et al. A blood-based prognostic biomarker in IBD. Gut 68, 1386–1395 (2019).

15. Thompson, B. Canonical Correlation Analysis: Uses and Interpretation. (SAGE, 1984).

16. Haghverdi, L., Lun, A. T. L., Morgan, M. D. & Marioni, J. C. Batch effects in single-cell RNA-sequencing data are corrected by matching mutual nearest neighbors. Nat. Biotechnol. 36, 421–427 (2018).

17. Rangel-Frausto, M. S. et al. The natural history of the systemic inflammatory response syndrome (SIRS). A prospective study. JAMA 273, 117–123 (1995).

18. Simpson, S. Q. SIRS in the Time of Sepsis-3. Chest 153, 34–38 (2018).

19. Rhee, C. & Klompas, M. Elucidating the Spectrum of Disease Severity Encompassed by Sepsis. JAMA Netw Open 5, e2147888 (2022).

20. Coifman, R. R. & Lafon, S. Diffusion maps. Appl. Comput. Harmon. Anal. 21, 5–30 (2006).

21. Pearl, J. Interpretation and Identification of Causal Mediation. Psychol. Methods 19, 459–481 (2014).

22. Imai, K., Keele, L. & Tingley, D. A general approach to causal mediation analysis. Psychol. Methods 15, 309–334 (2010).

23. Imai, K., Keele, L. & Yamamoto, T. Identification, Inference and Sensitivity Analysis for Causal Mediation Effects. SSO Schweiz. Monatsschr. Zahnheilkd. 25, 51–71 (2010).

24. Parnell, G. P. et al. Identifying key regulatory genes in the whole blood of septic patients to monitor underlying immune dysfunctions. Shock 40, 166–174 (2013).

25. Dunning, J. et al. Progression of whole-blood transcriptional signatures from interferon-induced to neutrophil-associated patterns in severe influenza. Nat. Immunol. 19, 625– 635 (2018).

26. COMBAT Consortium. A blood atlas of COVID-19 defines hallmarks of disease severity and specificity. Cell (2022) doi:10.1016/j.cell.2022.01.012.

27. Aschenbrenner, A. C. et al. Disease severity-specific neutrophil signatures in blood transcriptomes stratify COVID-19 patients. Genome Med. 13, 7 (2021).

28. Lee, J. C. et al. Gene expression profiling of CD8+ T cells predicts prognosis in patients with Crohn disease and ulcerative colitis. J. Clin. Invest. 121, 4170–4179 (10 2011).

29. Ulloa, L. & Tracey, K. J. The ‘cytokine profile’: a code for sepsis. Trends Mol. Med. 11, 56–63 (2005).

30. Singer, M. et al. The Third International Consensus Definitions for Sepsis and Septic Shock (Sepsis-3). JAMA 315, 801–810 (2016).

31. Xiao, W. et al. A genomic storm in critically injured humans. J. Exp. Med. 208, 2581– 2590 (2011).

32. Warren, H. S. et al. A genomic score prognostic of outcome in trauma patients. Mol. Med. 15, 220–227 (2009).

33. Zhang, Q. et al. Inborn errors of type I IFN immunity in patients with life-threatening COVID-19. Science 370, (2020).

34. Bastard, P. et al. Autoantibodies against type I IFNs in patients with life-threatening COVID-19. Science 370, (2020).

35. Hashiba, M. et al. Neutrophil extracellular traps in patients with sepsis. J. Surg. Res. 194, 248–254 (2015).

36. Mikacenic, C. et al. Neutrophil extracellular traps (NETs) are increased in the alveolar spaces of patients with ventilator-associated pneumonia. Crit. Care 22, 358 (2018).

37. Bone, R. C. et al. Definitions for sepsis and organ failure and guidelines for the use of innovative therapies in sepsis. The ACCP/SCCM Consensus Conference Committee. American College of Chest Physicians/Society of Critical Care Medicine. Chest 101, 1644–1655 (1992).

38. Huber, W., von Heydebreck, A., Sültmann, H., Poustka, A. & Vingron, M. Variance stabilization applied to microarray data calibration and to the quantification of differential expression. Bioinformatics 18, S96–S104 (2002).

39. Westra, H.-J. et al. MixupMapper: correcting sample mix-ups in genome-wide datasets increases power to detect small genetic effects. Bioinformatics 27, 2104–2111 (2011).

40. Johnson, W. E., Li, C. & Rabinovic, A. Adjusting batch effects in microarray expression data using empirical Bayes methods. Biostatistics 8, 118–127 (2006).

41. Dobin, A. et al. STAR: ultrafast universal RNA-seq aligner. Bioinformatics 29, 15–21 (2013).

42. Liao, Y., Smyth, G. K. & Shi, W. featureCounts: an efficient general purpose program for assigning sequence reads to genomic features. Bioinformatics 30, 923–930 (2014).

43. Thellin, O. et al. Housekeeping genes as internal standards: use and limits. J. Biotechnol. 75, 291–295 (1999).

44. Ritchie, M. E. et al. limma powers differential expression analyses for RNA-sequencing and microarray studies. Nucleic Acids Res. 43, e47 (2015).

45. Inouye, M. et al. An Immune Response Network Associated with Blood Lipid Levels. PLoS Genet. 6, e1001113 (2010).

46. Mayerle, J. et al. Identification of genetic loci associated with Helicobacter pylori serologic status. JAMA 309, 1912–1920 (2013).

47. Westra, H.-J. et al. Systematic identification of trans eQTLs as putative drivers of known disease associations. Nat. Genet. 45, 1238–1243 (2013).

48. Aguirre-Gamboa, R. et al. Deconvolution of bulk blood eQTL effects into immune cell subpopulations. BMC Bioinformatics 21, 243 (2020).

49. Netea, M. G. et al. Understanding human immune function using the resources from the Human Functional Genomics Project. Nat. Med. 22, 831–833 (2016).

50. COMBAT Consortium et al. A blood atlas of COVID-19 defines hallmarks of disease severity and specificity. bioRxiv (2021) doi:10.1101/2021.05.11.21256877.

51. Witten, D. M., Tibshirani, R. & Hastie, T. A penalized matrix decomposition, with applications to sparse principal components and canonical correlation analysis. Biostatistics 10, 515–534 (2009).

52. Angerer, P. et al. destiny: diffusion maps for large-scale single-cell data in R. Bioinformatics 32, 1241–1243 (2016).

53. Breiman, L. Random Forests. Mach. Learn. 45, 5–32 (2001).

54. Kuhn, M. caret: Classification and Regression Training. Astrophysics Source Code Library ascl:1505.003 (2015).

55. Liaw, A., Wiener, M. & Others. Classification and regression by randomForest. R news 2, 18–22 (2002).

56. Cohen, J. A Coefficient of Agreement for Nominal Scales. Educ. Psychol. Meas. 20, 37– 46 (1960).

57. Fang, H., Knezevic, B., Burnham, K. L. & Knight, J. C. XGR software for enhanced interpretation of genomic summary data, illustrated by application to immunological traits. Genome Med. 8, 129 (2016).

58. Jassal, B. et al. The reactome pathway knowledgebase. Nucleic Acids Res. 48, D498– D503 (2019).

59. Kruskal, W. H. & Wallis, W. A. Use of Ranks in One-Criterion Variance Analysis. J. Am. Stat. Assoc. 47, 583–621 (1952).

60. McCullagh, P. Regression Models for Ordinal Data. J. R. Stat. Soc. Series B Stat. Methodol. 42, 109–142 (1980).

61. Venables, W. N. & Ripley, B. D. Modern Applied Statistics with S. (Springer, New York, NY, 2002).

62. Therneau, T. M. & Grambsch, P. M. Modeling Survival Data: Extending the Cox Model. (Springer, New York, NY, 2000).

63. Zeileis, A. & Grothendieck, G. zoo: S3 Infrastructure for Regular and Irregular Time Series. Journal of Statistical Software, Articles 14, 1–27 (2005).

64. Cox, D. R. Regression models and life-tables. J. R. Stat. Soc. 34, 187–202 (1972).

65. Baron, R. M. & Kenny, D. A. The moderator-mediator variable distinction in social psychological research: conceptual, strategic, and statistical considerations. J. Pers. Soc. Psychol. 51, 1173–1182 (1986).

66. King, G., Tomz, M. & Wittenberg, J. Making the Most of Statistical Analyses: Improving Interpretation and Presentation. Am. J. Pol. Sci. 44, 347–361 (2000).

67. Tingley, D., Yamamoto, T., Hirose, K., Keele, L. & Imai, K. mediation: R package for causal mediation analysis. (2014).

